# Community structured model for vaccine strategies to control COVID19 spread: a mathematical study

**DOI:** 10.1101/2021.01.25.21250505

**Authors:** Elena Aruffo, Pei Yuan, Yi Tan, Evgenia Gatov, Effie Gournis, Sarah Collier, Nick Ogden, Jacques Bélair, Huaiping Zhu

**Affiliations:** Centre for Diseases Modeling (CDM), York University, Toronto, Ontario, Canada; Department of Mathematics and Statistics, York University, Toronto, Ontario, Canada; Toronto Public Health, Toronto, Ontario, Canada; Public Health Agency of Canada, Ottawa, Ontario, Canada; Département de Mathématiques et de Statistique, Université de Montréal, Montréal, Québec, Canada

## Abstract

Efforts to mitigate the COVID-19 pandemic have relied heavily on non-pharmaceutical interventions (*NPIs*), including physical distancing, hand hygiene, and mask-wearing. However, an effective vaccine is essential to containing the spread of the virus. The first doses were distributed at the end of 2020, but the efficacy, period of immunity it will provide, and percentage of coverage still remain unclear. We developed a compartment model to examine different vaccine strategies for controlling the spread of COVID-19. Our framework accounts for testing rates, test-turnaround times, and vaccination waning immunity. Using reported case data from the city of Toronto, Canada between Mar-Dec, 2020 we defined epidemic phases of infection using contact rates, which depend on individuals’ duration of time spent within the household, workplace/school, or community settings, as well as the probability of transmission upon contact. We investigated the impact of vaccine distribution by comparing different permutations of waning immunity, vaccine coverage and efficacy throughout various stages of NPI’s relaxation in terms of cases, deaths, and household transmission, as measured using the basic reproduction number (R_0_). We observed that widespread vaccine coverage substantially reduced the number of cases and deaths. In order for NPIs to be relaxed 8 months after vaccine distribution, infection spread can be kept under control with either 60% vaccine coverage, no waning immunity, and 70% efficacy, or with 60% coverage with a 12-month waning immunity and 90% vaccine efficacy. Widespread virus resurgence can result when the immunity wanes under 3 months and/or when NPI’s are relaxed in concomitance with vaccine distribution. In addition to vaccination, our analysis of R_0_ showed that the basic reproduction number is reduced by decreasing the tests turnaround time and transmission in the household. While we found that household transmission can decrease following the introduction of a vaccine, public health efforts to reduce test turnaround times remain important for virus containment. Our findings suggest that vaccinating two-thirds of the population with a vaccine that is at least 70% effective may be sufficient for controlling COVID-19 spread, as long as NPI’s are not immediately relaxed.

## Introduction

Since the World Health Organization declared the rapid spread of SARS-CoV-2 a global pandemic on March 11, 2020[1], initial control measures that are still in use in most affected countries are non-pharmaceutical interventions (*NPIs*, including physical distancing, hand hygiene, and mask-wearing), which have been efficient in controlling virus spread [2,3,4]. However, due to SARS-CoV-2 rapid spread and high mortality, it is widely accepted that a safe and effective vaccine is crucial for mitigating the epidemic [5,6] The scientific community has immediately started studying the virus and developing a vaccine against it. Several such vaccines are already available to multiple countries [7,8], while others are currently in clinical and preclinical development [9]. Given limited available doses and the increasing epidemic trend, a safe and effective vaccination strategy is paramount. Planning for the optimal distribution of the vaccine, and ultimately, determining whether and at what point the safe relaxation of NPIs is possible are critical public health objectives [5,6].

Several studies have quantified the effect of COVID-19 vaccination by using mathematical models [10,11,12,13,14,15,16,17]. For example, one study has utilised an information-dependent SIRI model and considered the occurrence of re-infection [14], showing that case counts can be reduced when vaccination is introduced into the population. However, latent stage, mortality, and implementation of quarantine/self-isolation were not considered. Another study has incorporated vaccination and isolation as key parameters in a SEIR model and compared the results of different vaccination coverage and isolation days [15]. While this study found that long isolation and high vaccine coverage can decrease virus spread, it did not account for hospitalizations, different vaccine efficacy levels, the degree of waning immunity, and the presence of asymptomatic infections, which are known to play an important role in spreading the virus [18,19]. Moreover, the specific context around vaccine implementation has not been investigated in depth. Many regions have escalated/de-escalated public health measures in timed stages to control virus spread [20,21,22]. The implementation of these policy phases depends on the local epidemic conditions and test processes. Accordingly, vaccine implementation should also consider the stages of the local epidemic trajectory. Lastly, despite the importance of detection of positive cases and test turnaround times [23], to our knowledge, few studies have considered testing processes in the context of vaccine modeling.

To fill these gaps, we aimed to develop an extensive compartmental model to investigate the impact of different scenarios of vaccine implementation on COVID-19 cases and deaths and the basic reproductive number. We simulate different degrees of waning immunity, vaccine efficacy, and population coverage, along with different time points at which NPIs are relaxed, in order to determine, under different conditions, the vaccine coverage needed to prevent a possible COVID-19 resurgence. To make our work more widely applicable to different regions, we consider different phases of the local epidemic trajectory and related public health mitigation strategies, as well as testing processes (including turnaround time).

## Materials and methods

### Compartmental model

We developed a Susceptible-Exposed-Infectious Asymptomatic-Infectious Mild-Infectious Quarantined-Hospitalized-Deceased-Recovered (*SEAI*_*m*_ *I*_*q*_ *HDR*) model with an infection-related death framework. We defined different phases using public health data related to policy (i.e., lockdowns, re-opening), test positivity, and test-turnaround times. To better reflect individuals’ activities as they relate to these phases, we divided the daily routine of the population into hours spent at home, in the community, or at work/school. Vaccination is incorporated by reducing the susceptible class and increasing the recovered class, accounting for various degrees of waning immunity. We investigated the impact of vaccination on cumulative cases and deaths, as well ass transmission (using the effective reproductive number) under different scenarios of vaccine coverage (10%, 30%, 60% or 90% of the population immunized via inoculation), vaccine efficacy [24] (70% or 90%), and waning immunity (none, 12 months, 3 months). These scenarios are simulated in conjunction with NPIs relaxation.

### Data source and phases

We developed a novel modeling framework that can be easily applied to any geographical context. As a case study, we used publicly available data on reported cases and deaths, test turnaround times in Toronto from March 17 to December 2, 2020, and percentage positivity test in Ontario, Canada [25,26] from April 19 17 to December 2, 2020, as well as information about the local epidemic trajectory and corresponding policy [27] around NPIs, school/business closures, testing processes and test turnaround times to define defined the following phases (Fig 1.):

**Fig 1.**
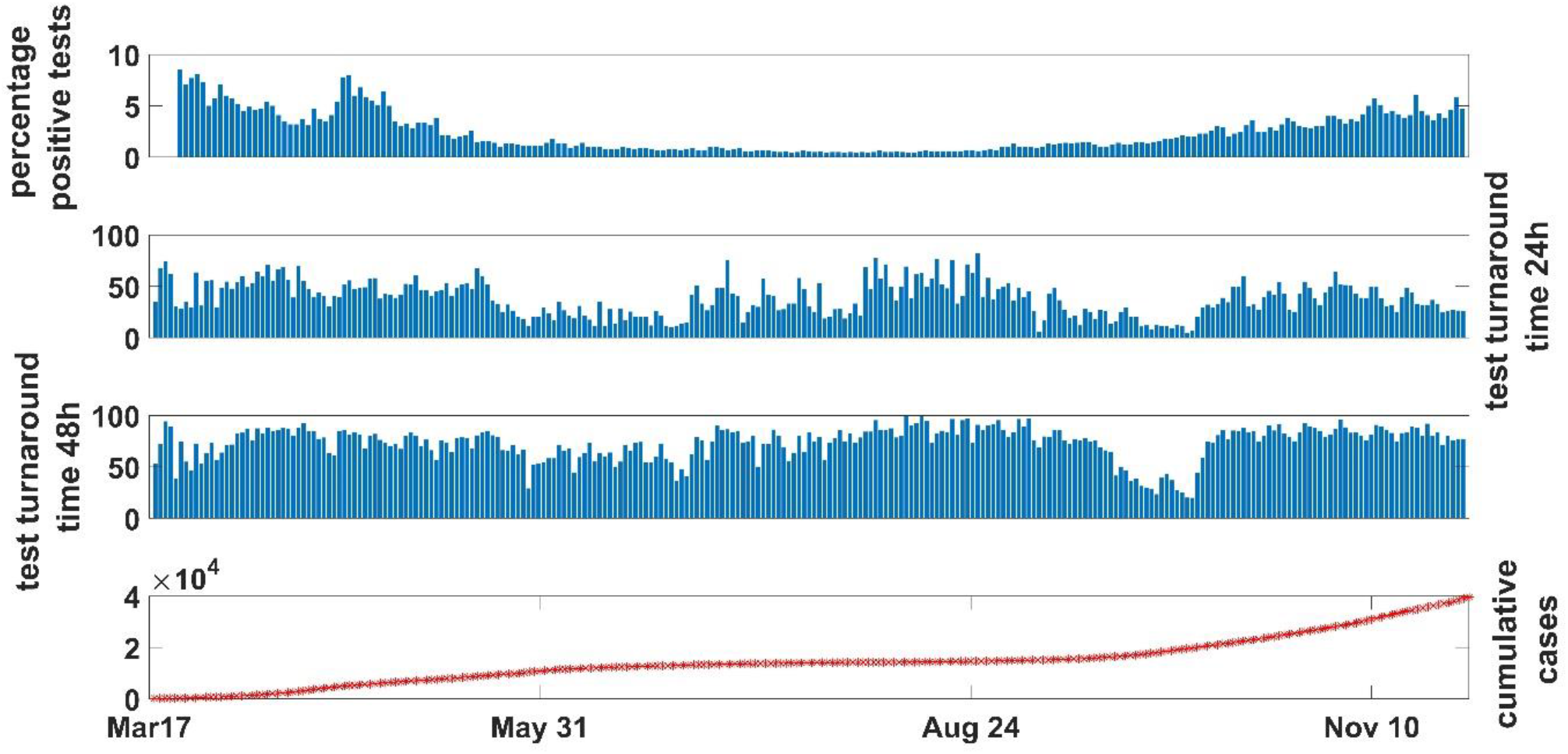
Percentage positive tests, tests turnaround times (24h-48h) and cumulative cases are reported from March 17, 2020 to November 28, 2020 in Toronto. The percentage positive tests decrease from May 31 to August 24, when it starts increasing, showing, however a magnitude smaller than the first wave in March. Until May 31 about 50% of the tests were returned within 24h, while until the end of August, the trend shows an initial decreasing followed by an increasing trend. After this, the curve decreases again, raising up in October and remaining below 50%. However, it slightly decreases again after November 10. Similar pattern for 48h tests turnaround time. The cumulative cases increase until May 31, and then remain stable until the end of August when the increase starts again and continue until Dec 2, 2020, with a slight decrease after the beginning of November.

1. March 17 (April 19)-May 31 (Lockdown): Schools and public spaces are closed, resulting in limited time and contact in the community, with somewhat lenient NPIs. Testing is not widespread, with 50% of results returned within 24 hours and percent positivity between 8-10%. There was a slight increase in cumulative cases.
2. June 1-August 24 (Re-opening stages): Businesses gradually open, resulting in increased contact and time spent in the community, and stricter NPIs. Widespread testing is in place, with 24h turnaround under 50% initially, then rising in the summer months. Percent positivity dropped to 1.2%. Infections seem to have stabilized.
3. August 25-November 10 (Resurgence starts): Schools fully reopened in September, followed by escalating restrictions in October and November (e.g., closure of indoor dining, restriction on the number of people indoors), resulting in decreasing time in the community under strict NPIs. Testing is widespread, with less than 50% returned within 24 hours, and a 5% percent positivity. Case counts begin to rise more steeply, compared to before the March lockdown.
4. November 10-December 2 (Additional closures): Schools are open, with additional closures of public spaces. The public is encouraged to stay at home. Test turnaround within 24h is < 50%, with a 5% percent positivity. The sharp rise in cases stabilizes slightly.

### Community structured transmission model

Using the least square method, we fit our model to estimate parameters for each phase. We propose a model following a Susceptible-Exposed-Infectious Asymptomatic-Infectious Mild-Infectious Quarantined-Hospitalized-Deceased-Recovered (*SEI*_*A*_ *I*_*m*_ *I*_*q*_ *HDR*) framework, where demographics are ignored and death due to infection, is included (Table 1).

**Table 1:**
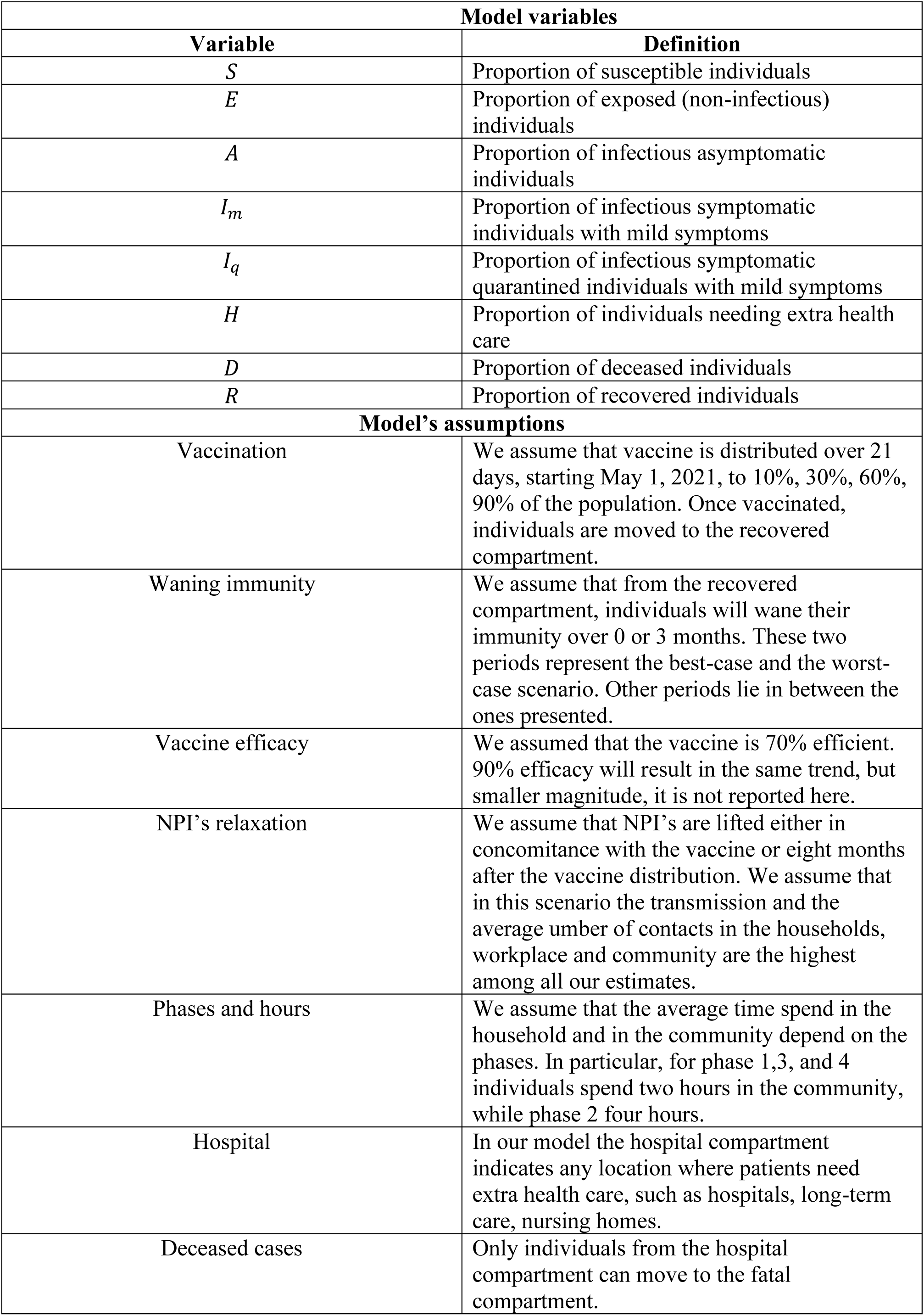
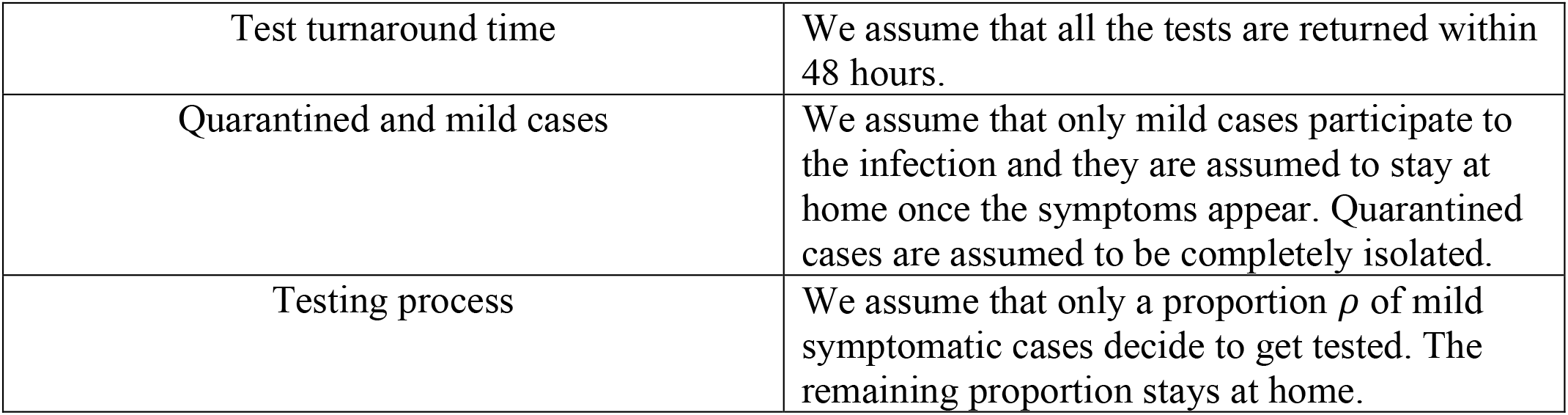
Model variables and assumptions

| Table 1. Model variables |  |
| --- | --- |
| Variable | Definition |
| $S$ | Proportion of susceptible individuals |
| $E$ | Proportion of exposed (non-infectious) individuals |
| $A$ | Proportion of infectious asymptomatic individuals |
| $I_m$ | Proportion of infectious symptomatic individuals with mild symptoms |
| $I_q$ | Proportion of infectious symptomatic quarantined individuals with mild symptoms |
| $H$ | Proportion of individuals needing extra health care |
| $D$ | Proportion of deceased individuals |
| $R$ | Proportion of recovered individuals |
| Model's assumptions |  |
| Vaccination | We assume that vaccine is distributed over 21 days, starting May 1, 2021, to 10%, 30%, 60%, 90% of the population. Once vaccinated, individuals are moved to the recovered compartment. |
| Waning immunity | We assume that from the recovered compartment, individuals will wane their immunity over 0 or 3 months. These two periods represent the best-case and the worst-case scenario. Other periods lie in between the ones presented. |
| Vaccine efficacy | We assumed that the vaccine is 70% efficient. 90% efficacy will result in the same trend, but smaller magnitude, it is not reported here. |
| NPI's relaxation | We assume that NPI's are lifted either in concomitance with the vaccine or eight months after the vaccine distribution. We assume that in this scenario the transmission and the average number of contacts in the households, workplace and community are the highest among all our estimates. |
| Phases and hours | We assume that the average time spend in the household and in the community depend on the phases. In particular, for phase 1,3, and 4 individuals spend two hours in the community, while phase 2 four hours. |
| Hospital | In our model the hospital compartment indicates any location where patients need extra health care, such as hospitals, long-term care, nursing homes. |
| Deceased cases | Only individuals from the hospital compartment can move to the fatal compartment. |
| Test turnaround time | We assume that all the tests are returned within 48 hours. |
| Quarantined and mild cases | We assume that only mild cases participate to the infection and they are assumed to stay at home once the symptoms appear. Quarantined cases are assumed to be completely isolated. |
| Testing process | We assume that only a proportion $\rho$ of mild symptomatic cases decide to get tested. The remaining proportion stays at home. |

The vaccination process is included in the model by reducing the susceptible class and moving immunized individuals into the recovered compartment (Fig 2.). We partition the day based on the daily activities in the work/home, community, or home settings. Each classification is represented by its own probability of transmission term and average contact rate.

**Fig 2.**
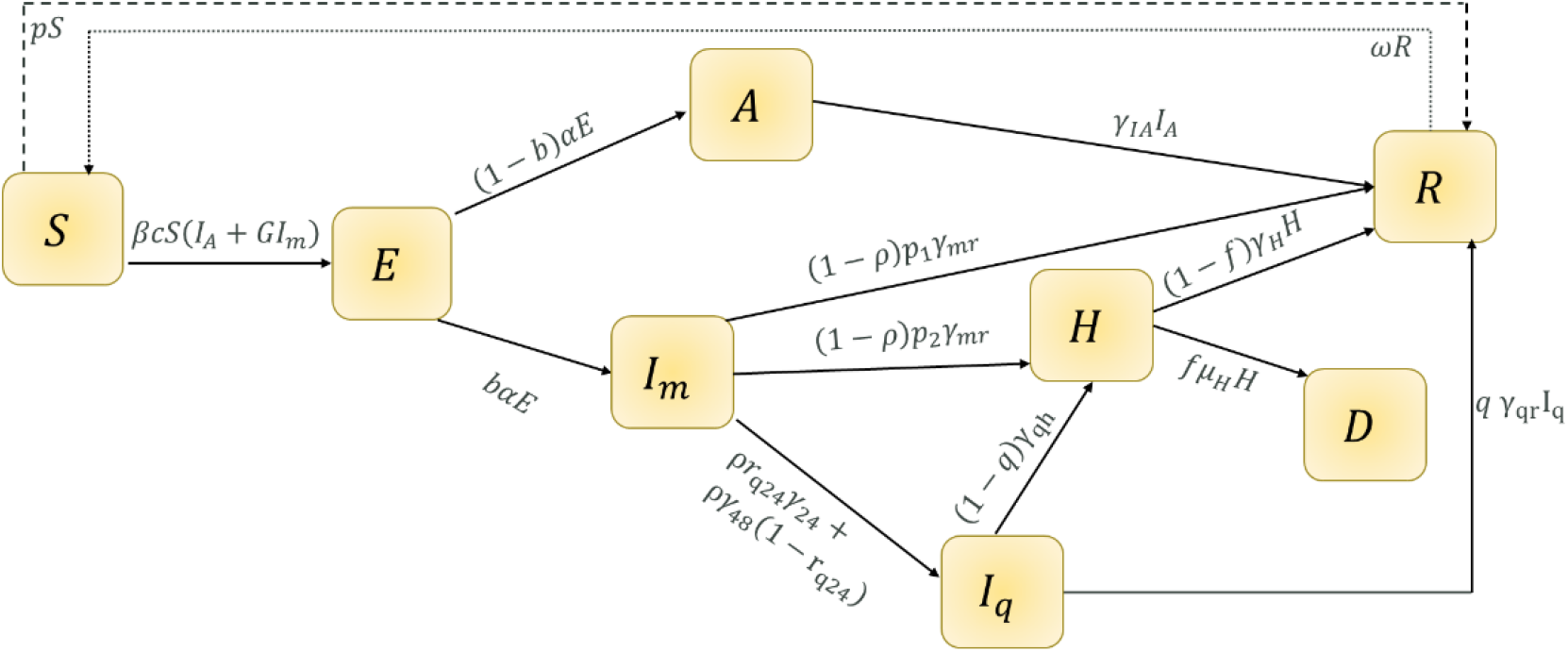
Flow diagram of Model (1). The vaccine process is defined with dashed lines while the waning immunity process is indicated with dotted lines.

COVID-19 transmission occurs as follows: susceptible individuals become infected after encountering infectious asymptomatic and mild symptomatic cases. However, since mild cases are assumed to stay at home, they contribute only to household transmission. Quarantined cases are assumed to have no contact; hence they do not transmit the infection. Mild and quarantined cases might develop severe symptoms and become hospitalized or recover from mild status. We assume that only hospitalized patients might die from infection. We also included the testing process, assuming that a proportion *ρ* of mild symptomatic cases get tested. Once tested, some (*r*_24_) will receive results within 24 hours and the rest will receive results within 48 hours. Once the vaccine is introduced into the population, immunity through inoculation might wane over time, making individuals susceptible again.

The general system of ODEs describing COVID-19 infection dynamics is given by (1).

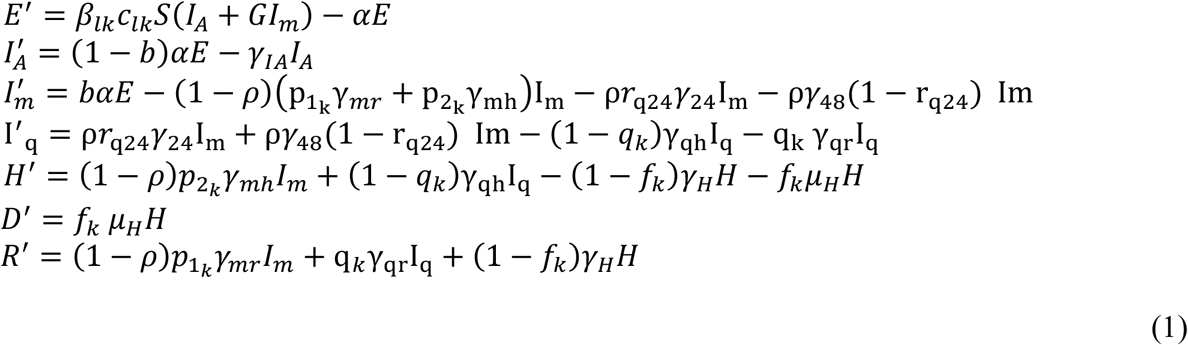

where *l* indicates the different locations and *k* indicated the different phases. A list of the parameters is provided in S1Table 1.

Given the large number of doses of vaccine planned to be distributed in the first stage of its implementation^24^, it is unrealistic that the distribution of doses occurs in a single day. Hence, we assume that *τ* is a random variable that describes the time during which the vaccine is implemented. Hence, *τ* follows an Exponential distribution

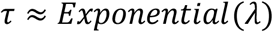

With 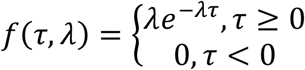, where *λ* is the daily average number of individuals in the population getting vaccinated. We can state that expectation 𝔼(*τ*) = 1/*λ* is the average time to vaccinate a certain proportion of population and the probability density function *f*(*τ, λ*) is the probability that an individual will be vaccinated in *τ* days. After the vaccine is introduced in the population, the waning process is included in the model.

Lastly, *p* is the proportion of the population that needs to be vaccinated and 1/*ω* is the waning period. Thus, we can modify the susceptible and recovered equation in system (1) as follows:

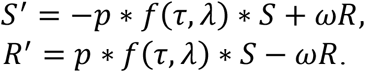

### Impact of vaccine introduction on COVID-19 trajectory

To study the impact of a vaccine and its biological processes on case numbers and deaths under different phases of the infection and different conditions, we provide a comprehensive analysis varying all these components. We compare different vaccine coverage (10%, 30%, 60%, 90%), assuming the doses are given over three weeks. Given the vaccine efficacy uncertainty, we propose projections when this factor is 70% or 90%. Also, we investigate the effect of waning immunity comparing the scenarios in which waning immunity never occurs or occurs over three or 12 months. Given that NPI’s have been playing an important role in controlling the epidemic, we analyze the impact of early (simultaneous with vaccine introduction) versus late (8 months subsequently) relaxation of NPIs, assuming that the probabilities of transmission and the average number of contacts are the highest among all the phases, after vaccine is implemented.

We investigate the basic reproduction number by introducing four-dimensional contour plots. These plots allow us to investigate how the reproduction number is changing when three parameters are varied over given intervals and immunity does not wane. This provides a comprehensive understanding of the relationship between *R*_0_ and the parameters in our analysis. We examined the basic reproduction number ℛ_0_ using the next-generation matrix method (details in Supporting Information 2). We derived the following expression:

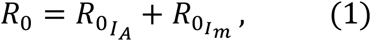

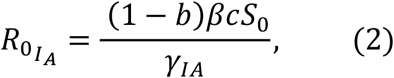

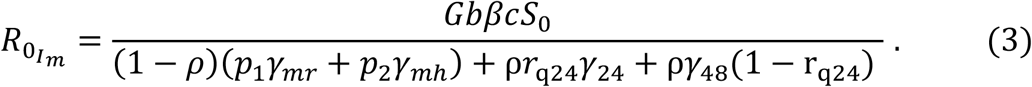

Eq. (1) shows the contribution that asymptomatic and mild cases have on generating new infections. Since mild cases are always at home, this expression highlights the contribution that the household transmission has on the spread of the virus.

## Results

### Reproduction number

Fig 3. illustrates 4D contour plots of the household reproduction number in the *β*_*h*_ − *r*_24_ − *ρ* parametric space. We present the scenario with no vaccination and with 30%, 90% vaccine coverage, 70% efficacy and lifelong immunity. *R*_0_ is less than 1 when the vaccine coverage is 90%. With vaccine efficacy of 90%, in Phases 1, 3 and 4 the vaccination coverage needed to provide a R_0_ less than one is 60%, while for Phase 2 it is 30% (see, Supporting Information S2.1 Fig). In Phase 3, characterized by the highest transmission, the R_0_ can get as high as 2, while, in Phase 2, characterized by the lowest transmission, the R_0_ reaches approximately 1.6 at most. An increase in vaccination coverage from 30% to 90% will decrease the R_0_ by 50%. The region indicating the highest values of *R*_0_ (yellow area) increases as houdrholf transmission increases. The blue area (for smaller *R*_0_) increases if more positive tests are detected and more tests are returned within 24 hours (i.e., *ρ* and *r*_24_ increase).

**Fig 3.**
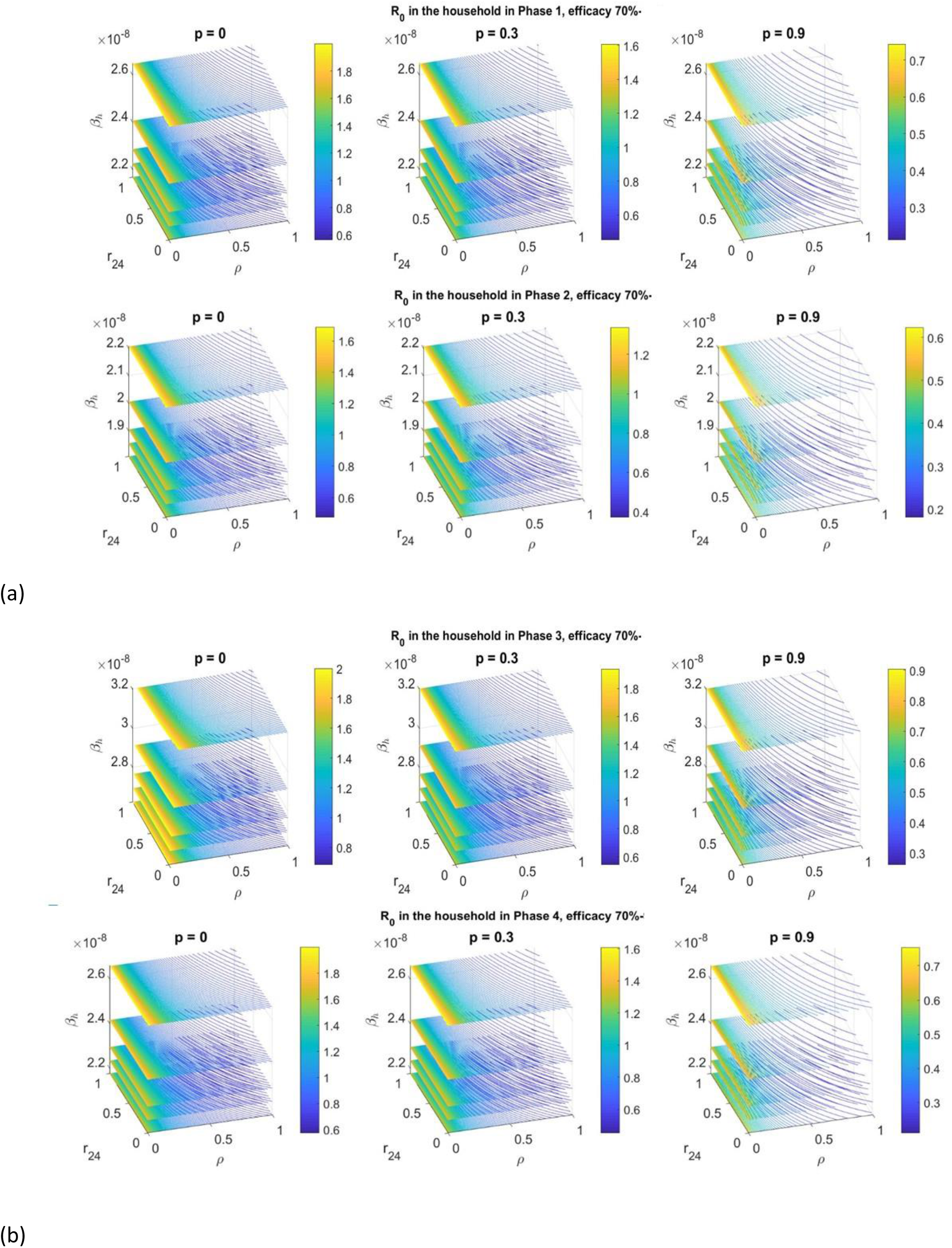
*R*_0_ 4D contour plot in the *β*_*h*_, *r*_24_, *ρ* parameters space. The values of the reproduction number are presented under Phase 1-2 and 3-4 (a, b, respectively) considering 0%, 30%,90% vaccine coverage, lifelong immunity and efficacy 70%. In all cases, we observe that as the vaccine coverage increases, the reproduction number decreases visibly. Also, the yellow region (indicating the highest values of the reproduction number) increases as the probability of the transmission in the household increases. We observe that if less than 1% of mild cases is tested, the reproduction number becomes larger. The more mild cases are tests, the smaller the reproduction number. Also, it is visible that to reduce the infection’s spread, more people need to receive their test within 24h.

### Long term projections: cases and deaths

In this section, we report the dynamic of the infection if immunity does not wane or wanes over 3 months. The trend of decreasing case numbers is very similar to the one shown by the cumulative cases; hence they are reported in the Supporting Information (S3.2-3.5-3.8-3.11 Figs.). The trend of the infection in Phase 1,2 and 4 are similar, so the figure for the latter cases are reported in the Supporting Information (S3.4-S3.10 Figs). We present scenarios whereby a 70% effective vaccine, is introduced in the population on May 1, 2021 (coverage: 10%, orange, 30%, purple, 60% light blue, 90% blue). NPI’s relaxed either in concomitance with vaccine introduction or 8 months subsequently. The results of cumulative cases when efficacy is 90% are reported in Supporting Information (S3.3-3.6-3.9-3.12 Figs).

### Prediction under Phase 1, 2 and 4

The infection trend under Phase 1, 2, and 4 appears similar (Figs 4, S3.4, S3.10). In general, higher vaccine coverage saw decreasing infections. Vaccine-induced immunity waning over 3 months (Fig 3A.) results in the re-emergence of the infection, more so when NPI’s are lifted early. If immunity does not wane and NPI’s are lifted early (Fig 4B. left), infection resurgence is observed even if 60% of the population is vaccinated. By contrast, if NPIs are relaxed later (Fig 4B. right), 60% coverage is enough to control the epidemic and prevent resurgence. In all scenarios, neither 10% nor 30% population coverage ensures long-term control of the infection. With 12-month waning immunity (Fig S3.1C), outcomes are similar to 3-month waning, though with a smaller magnitude. A 90% efficient vaccine shows a similar trend to the 70% effective vaccine, tough with smaller magnitude (Fig S3.3). In all scenarios, we observed that postponing the relaxation of NPIs, compared to early relaxation, results in roughly a 30% decrease in cases and deaths by June 2022.

**Fig 4.**
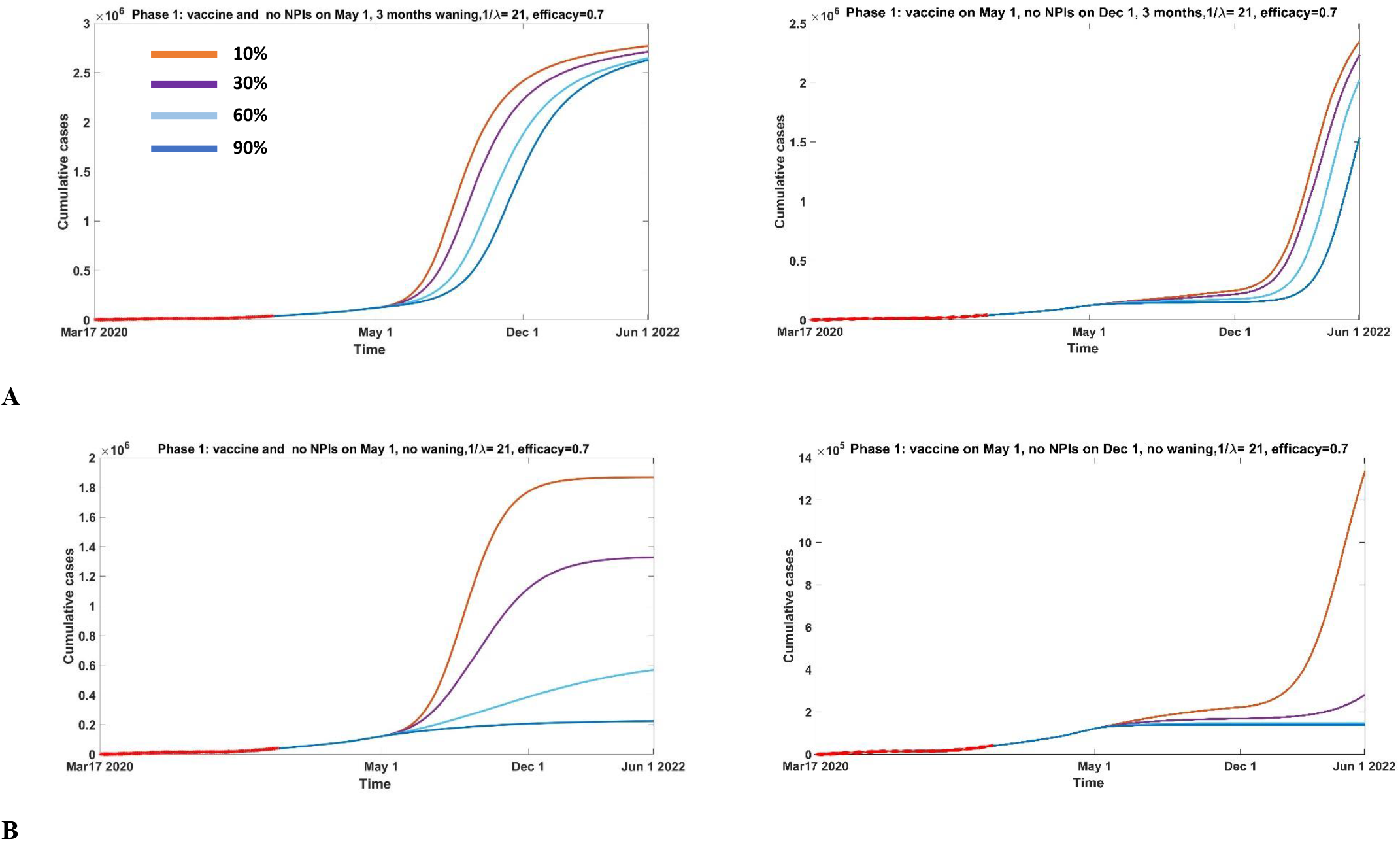
Cumulative cases from March 17, 2020 to June 2022 under Phase 1. Vaccine is introduced on May 1, 2021 (10%, orange, 30%, purple, 60% light blue, 90% blue) and distributed over 21 days. NPI’s are lifted (highest number of contacts and probability of transmission) on December 1, 2021 (A-B right panels) or in concomitance with vaccine (A-B left panels), immunity wanes over 3 months (A) or does not wane (B), and vaccine is 70% efficient.

### Prediction under Phase 3

In Phase 3 scenario (Fig 5), vaccination reduces the spread of the infection. We immediately observe that when immunity wanes quickly, and NPIs are lifted later (Fig 5A right), the trend shows a slight increase after the vaccine is implemented and a sharper increase once NPIs are lifted. Earlier relaxation of NPIs (Fig 5A left) will result in an immediate increase in cases, peaking before September (or December, depending on the vaccine coverage) without remerging afterward. Although the trend changes between the two NPI’s relaxation scenarios, the magnitude of cases and deaths does not seem to vary considerably. However, if NPIs are lifted later, 90% of population coverage results in no new cases, while 60% coverage results in a slight increase. If immunity does not wane (Fig 5B), cumulative cases reach a maximum magnitude and then stabilize. If the NPIs are relaxed later (Fig 5B right), the trend does not change remarkably for 60% or 90% coverage. In Phase 3, since initial transmission is very high, postponed relaxation of NPIs does not result in a sufficient decrease in cases.

**Fig 5.**
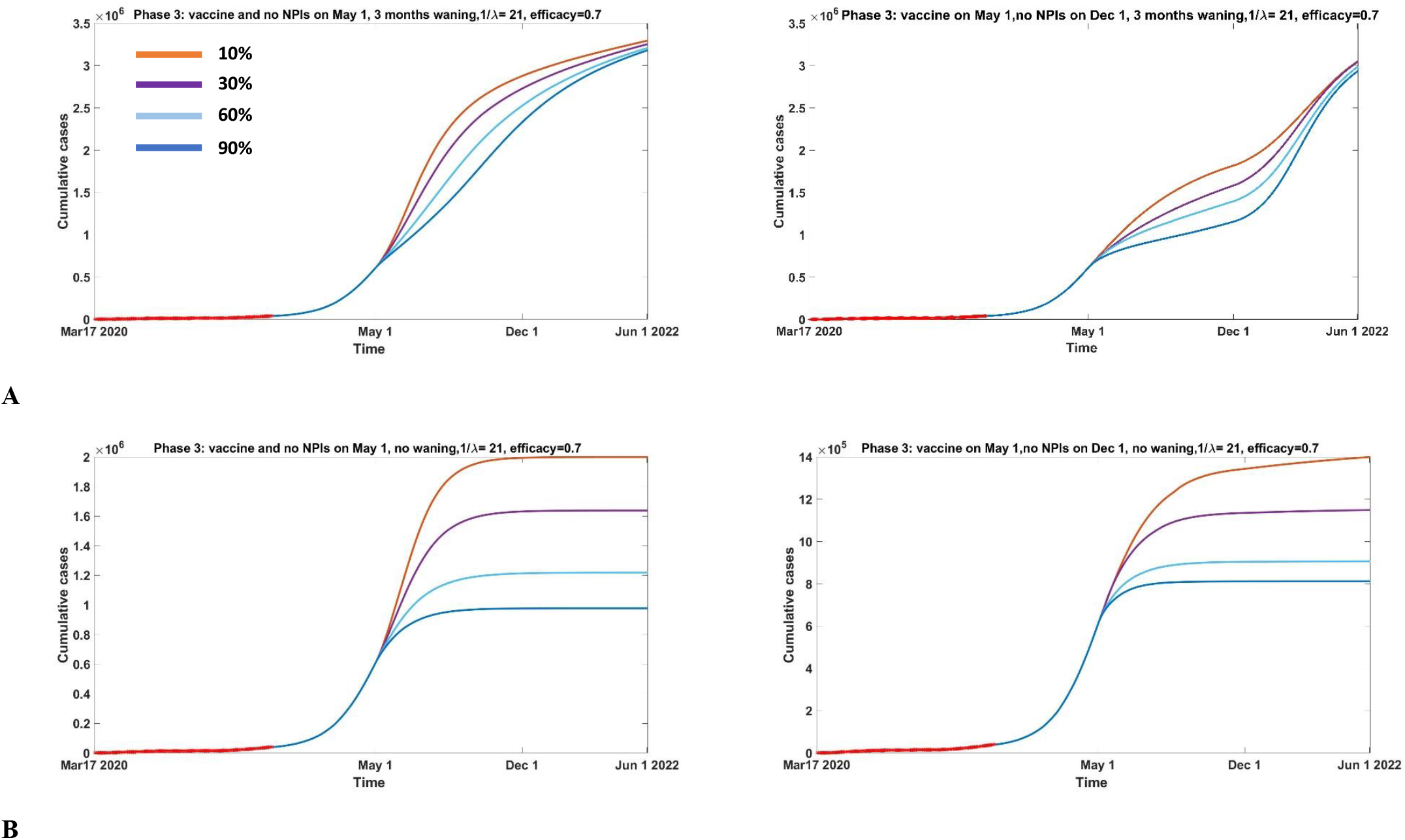
Cumulative cases from March 17, 2020 to June 2022 under Phase 3. Vaccine is introduced on May 1, 2021 (10%, orange, 30%, purple, 60% light blue, 90% blue) and distributed over 21 days. NPI’s are lifted (highest number of contacts and probability of transmission) on December 1, 2021 (A-B right panels) or in concomitance with vaccine (A-B left panels), immunity wanes over 3 months (A) or does not wane (B), and vaccine is 70% efficient.

## Conclusions

We have introduced a compartmental model to examine a global vaccination strategy to control COVID-19 outbreaks given different scenarios of the vaccine’s coverage and effectiveness, and waning immunity. We considered different time points of NPI relaxation and took into account different phases of the epidemic trajectory and testing processes. By investigating testing rates, turnaround times, and the restrictions enforced by public health, we were able to provide objective criteria to describe epidemic’s trajectory through distinct phases. We use Toronto as case study. Our model does not directly describe an aged-structured population, but rather incorporates the activity level of the population considering different transmission and contact rates that depend on the hourly distribution, on a daily basis, of the time spent at work/school, in the community, and at home. Since activity in these settings is impacted by public health policy, this differentiation helps us understand which location drives the transmission, as measured by the reproductive number. Our data fitting shows that household transmission is much higher than transmission at work/school or community, even though the number of contacts is smaller: this can undoubtedly be attributed to the *duration* of contacts within households. Therefore, it is crucial that public health focuses on reducing transmission in the household, by promoting sanitizing and hand-hygiene processes: we suspect that, in general, less attention is paid to these measures *within* households. As expected, the introduction of vaccination reduces transmission. Moreover, to keep epidemic spread under control, increasing the number of positive tests returned within 24 hours is paramount. This finding suggests a need for increasing public health resources for the quick identification of COVID-19 positive cases.

We generated forecasts of cumulative cases and deaths following vaccine implantation, considering different levels of efficacy (70%-90%) and different degrees of waning immunity (0, 3, 12 months). Vaccination follows an exponential distribution, assuming distribution over a three-week period. Our results suggest that rapid waning immunity, independently from vaccine coverage, will result in disease resurgence. With lifelong immunity, vaccine coverage above 60% is sufficient to prevent re-emergence of the outbreak. Under epidemic conditions characterised by fewer public health restrictions and increased contact rates in the community, early relaxation of NPIs will result in a 30% increase in cases and deaths, compared to late relaxation of NPIs. However, in the lowest transmission scenario, corresponding to a timeframe characterised by lockdowns and low contact rates, even with a 3-month waning immunity, postponing NPI relaxation can result in decreases in cases and deaths, even more so with lifelong immunity. This suggests that under low transmission, and a large proportion of susceptibles, lifting NPIs will result in a rapid infection resurgence. Furthermore, in all scenarios, a 90% vaccine effectiveness results in further reductions in cases and deaths. Hence, we conclude that short immunity and early relaxation of NPIs are key drivers for disease resurgence. Future work is needed to further understand the role of waning processes and vaccine efficacy.

Our model has some limitations. Firstly, we consider the population as a whole. However, age and socioeconomic status are imperative to incorporate when vaccine policies need to be implemented. For example, in many countries that have started vaccinating, older individuals and those working in healthcare and other essential occupations have been prioritized in receiving the first doses. We are currently working on extending our work to include multiple age groups. Secondly, we assume that all mild cases stay at home, following a strict self-isolation once symptoms show. However, this might not be realistic, as the compliance to self-isolation protocols seem to be variable: this aspect will be included in future work. Thirdly, we assume that only one dose vaccine is provided. However, at the time of submission, it is evident that a second dose needs to be included to achieve full immunity.

Additionally, we assume that the hospital compartment includes all the structures where patients need extra health care, such as long-term care and hospitals, and also that deceased cases are only reported from this whole structure. However, the differentiation among all these structures needs to be included to understand better where public health needs to focus its resources. Similarly, we are ignoring internal hospital structures so that care facilities are treated as a single compartment: as some COVID-19 patients need a level of care that is treated in ICUs, a differentiation among severe cases in nosocomial needs to be included to better describe the hospitalized cases.

In conclusion, our methods, modeling the virus trajectory throughout different phases using epidemiological data combined with information on government restriction policies, make our results applicable to any geographic context for which sufficiently granular data are available. In fact, to understand the infection dynamics and the impact vaccination will have on it, it is crucial, but also sufficient, to identify the phase in the outbreak that a specific region is experiencing. Our findings that public health authorities emphasize the need for the population to pay attention to the transmission *inside* households, by urging strict compliance with sanitation measures upon arrival at home (wash hands, accessories, change of clothes, etc…). In order to prevent COVID-19 resurgence, higher population coverage is essential, along with a late relaxation of NPIs. Future work to determine the window of immunity following vaccination is needed to further refine our understanding of the required vaccine coverage to keep virus spread under control.

## Data Availability

the data is available in the manuscript and supporting information.

