## Supporting Information for "Community structured model for vaccine strategies to control COVID19 spread: a mathematical study"

**Affiliations:**

### Supporting Information 1

| Table 1. Model parameters | | | |
|---|---|---|---|
| Parameter | Definition | Value | Ref |
| $\beta_{h1}$ | Probability of transmission in the house PHASE1 | 2.4177e-08 | Estimated |
| $\beta_{c1}$ | Probability of transmission in the community PHASE1 | 3.3833e-09 | Estimated |
| $\beta_{h2}$ | Probability of transmission in the house PHASE2 | 1.5507e-08 | Estimated |
| $\beta_{c2}$ | Probability of transmission in the house community PHASE2 | 2.0633e-09 | Estimated |
| $\beta_{h3}$ | Probability of transmission in the house PHASE3 | 2.8142e-08 | Estimated |
| $\beta_{c3}$ | Probability of transmission in the community PHASE3 | 2.9199e-09 | Estimated |
| $\beta_{h4}$ | Probability of transmission in the house PHASE4 | 2.1143e-08 | Estimated |
| $\beta_{c4}$ | Probability of transmission in the community PHASE4 | 2.1000e-09 | Estimated |
| $c_h$ | Average daily contact at home | 2.4 | [1] |
| $c_{c_k}$ | Average daily contact in the community ($k = 1,2,3,4$ indicating the phases of the infection) | $c_{c_1}$ =6.4031<br>$c_{c_2}$ =11.4030<br>$c_{c_3}$ =11.5031<br>$c_{c_4}$ =6.4031 | Estimated |
| $c_{w_k}$ | Average daily contact in the community/work ($k = 1,2,3,4$ indicating the phases of the infection) | $c_{w_1}$ =8.4031<br>$c_{w_2}$ =11.5031<br>$c_{w_3}$ =11.5305<br>$c_{w_4}$ =10.1031 | Estimated |
| $\alpha$ | Average latent period | 1/4 $days^{-1}$ | [2,3] |
| $b$ | Proportion of symptomatic cases | 0.8 | [4] |
| $\rho_k$ | Testing rate ($k = 1,2,3,4$ indicating the phases of the infection) | $\rho_1$ =0.05999<br>$\rho_2$ =0.0100<br>$\rho_3 = 0.0399$<br>$\rho_4$ =0.0450 | Estimated |
| $p_{1_k}$ | Proportion of mild cases not tested who will recover as mild ($k = 1,2,3,4$ indicating the phases of the infection) | $p_{1_1}$ =0.7000<br>$p_{1_2} = 0.8000$<br>$p_{1_3} = 0.9000$<br>$p_{1_4} = 0.9000$ | Estimated |

| | | | |
|---|---|---|---|
| $p_{2_k}$ | Proportion of mild cases not tested who will become hospitalized ($k = 1,2,3,4$ indicating the phases of the infection) | $1 - p_{1_k}$ | Estimated |
| $\gamma_{mr}$ | Recovery rate of mild cases, not tested | 1/14 $days^{-1}$ | [5] |
| $\gamma_{qr}$ | Recovery rate of quarantined cases | 1/14 $days^{-1}$ | Assumed as $\gamma_{mr}$ |
| $\gamma_{mh}$ | Hospitalization rate of non-tested mild cases | 1/6 days^-1 | [6], assumed |
| $1/\gamma_{24,48}$ | Time needed to return tests | 24 hours, 48 hours | [7], assumed |
| $r_{24}$ | Proportion of individuals receiving their test result within 24 hours | $r_{24}$=0.5;<br>$r_{24}$0.45;<br>$r_{24}$=0.37;<br>$r_{24}$=0.37; | [7], assumed |
| $\gamma_{qh}$ | Hospitalization rate of quarantined cases | $\gamma_{qh_{1,2,3,4}}$ =0.25 | Estimated |
| $\gamma_H$ | Recovery rate of hospitalized | 1/10 $days^{-1}$ | [6], assumed |
| $q_k$ | Proportion of mild cases recovered ($k = 1,2,3,4$ indicating the phases of the infection) | $q_{1,2,3,4} = 0.7000$ | Estimated |
| $1 - q_k$ | Proportion of mild cases recovered ($k = 1,2,3,4$ indicating the phases of the infection) | | Estimated |
| $f_k$ | Proportion of hospitalized cases deceased ($k = 1,2,3,4$ indicating the phases of the infection) | $f_1 = 0.1999$<br>$f_2 = 0.070$<br>$f_3 = 0.070$<br>$f_4 = 0.059$ | Estimated |
| $1 - f_k$ | Proportion of hospitalized cases recovered ($k = 1,2,3,4$ indicating the phases of the infection) | | Estimated |
| $\omega$ | Waning periods | No waning, 3, 6, 12 months | Assumed |
| $p$ | Vaccination coverage | 10%, 30%, 60%, 90% | Assumed |
| $\mu_{H_k}$ | Mortality rate ($k = 1,2,3,4$ indicating the phases of the infection) | $\mu_{H_1} = 0.2499$<br>$\mu_{H_{2,3,4}}$ =0.1667 | Estimated |
| $G$ | Switch parameter (G=1 home hours- G=0 community hours) | G=1 home hours<br>G=0 community hours | Assumed |
| $E_0$ | Initial values of exposed individuals | 1.1000e+03 | Estimated |
| $A_0$ | Initial values of asymptomatic individuals | 130.0000 | Estimated |

| | | | |
|---|---|---|---|
| $I_{q_0}$ | Initial values of quarantined individuals | 10.0000 | Estimated |
| $I_{m_0}$ | Initial values of mild individuals | 133 | [8] |
| $H_o$ | Initial values of hospitalized cases | 32 | [8] |
| $D_0$ | Initial values deceased cases | 0 | [8] |
| $S_0$ | Initial values susceptible individuals | $2956024 - \mathrm{E}_0 - A_0 - I_{m_0} - I_{q_0} - R_0 - H_0 - D_0$ | calculated from [9] |

### Supporting Information 2: Reproduction numbers

Given System (1), we derive an expression for the reproduction number employing the next generation matrix method [10,11,12]. We start defining the matrices $F$ and $V$:

$$F = \begin{bmatrix} 0 & \beta c\, S_0 & \beta c S_0 & 0 \\ 0 & 0 & 0 & 0 \\ 0 & 0 & 0 & 0 \\ 0 & 0 & 0 & 0 \end{bmatrix}$$

$$-V = \begin{bmatrix} \alpha & 0 & 0 & 0 \\ -(1-b)\alpha & \gamma_{IA} & 0 & 0 \\ -b\alpha & 0 & (1-\rho)(p_1\gamma_{mr} + p_2\gamma_{mh}) + \rho r_{q24}\gamma_{24} + \rho\gamma_{48}(1 - \mathrm{r}_{q24}) & 0 \\ 0 & 0 & -\rho r_{q24}\gamma_{24} - \rho\gamma_{48}(1 - \mathrm{r}_{q24}) & -(1-\rho)(p_1\gamma_{mr} + p_2\gamma_{mh}) \end{bmatrix}$$

After evaluating the product matrix $F(-V)^{-1}$, the basic reproduction number is given by:

$$R_0 = \rho(F(-V)^{-1})$$

Where $\rho$ is the spectral radius of the matrix. The $R_0$ expression will then be:

$$R_0 = \beta c S_0 \left( \frac{1-b}{\gamma_{IA}} + \frac{Gb}{(1-\rho)(p_1\gamma_{mr} + p_2\gamma_{mh}) + \rho r_{q24}\gamma_{24} + \rho\gamma_{48}(1 - \mathrm{r}_{q24})} \right) \; (SM2.1)$$

If we observe this expression, we can express it in terms of the type reproduction numbers:

$$R_{0_{I_A}} = \frac{(1-b)\beta c S_0}{\gamma_{IA}} \; (SM2.2)$$

$$R_{0_{I_m}} = \frac{Gb\beta c S_0}{(1-\rho)(p_1\gamma_{mr} + p_2\gamma_{mh}) + \rho r_{q24}\gamma_{24} + \rho\gamma_{48}(1 - \mathrm{r}_{q24})} \; (SM2.3)$$

Hence

$$R_0 = R_{0_{I_A}} + R_{0_{I_m}} (SM3.4)$$

Eq. (SM2.1) provides us not just the contribution of asymptomatic and mild cases on the spread of the infection, it does also highlight the contribution that the household transmission and community transmission, at work or in the community, have on the progress of the outbreak. Also, we observe that if all mild cases test, then the number of new cases is strictly related to the time needed to get the results back. If the tests are returned fast enough ($\frac{1}{\gamma_{24}}$ *and* $\frac{1}{\gamma_{48}}$ *approach* 0 ), $R_{0_{I_m}} = 0$, and hence the basic reproduction number will just depend on the asymptomatic cases. Same result is obtained if a larger number of individuals receive the test within 24 hours.

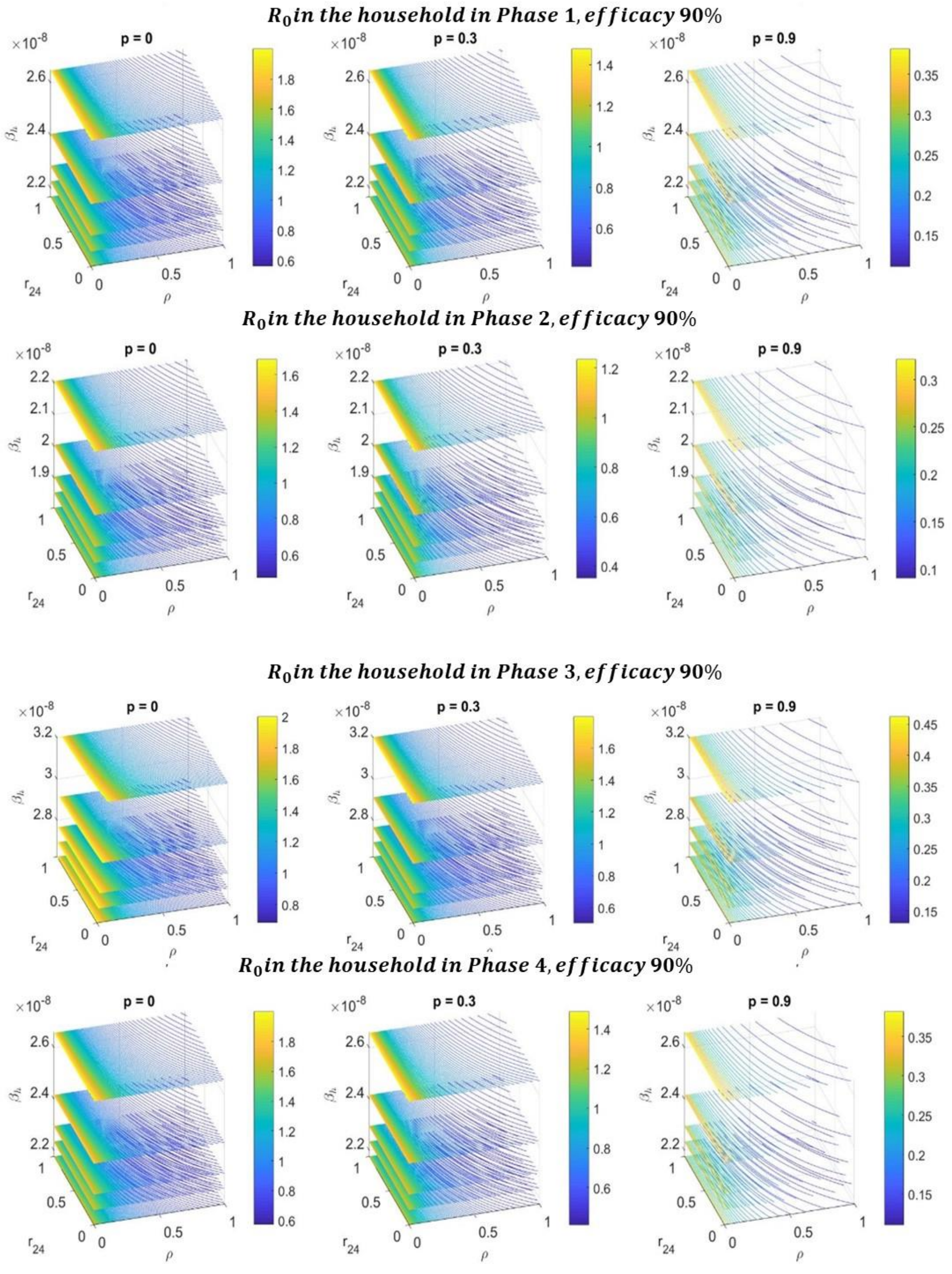


**Figure S2.1: $R_0$ 4D contour plot in the $\beta_h, r_{24}, \rho$ parameters space**. The values of the reproduction number are presented under Phase (A) 1 and 2 (B) 3 and 4 considering 0%, 30%,90% vaccine coverage, lifelong immunity and efficacy 90%

#### Supporting Information 3: Complete predictions

##### Phase 1

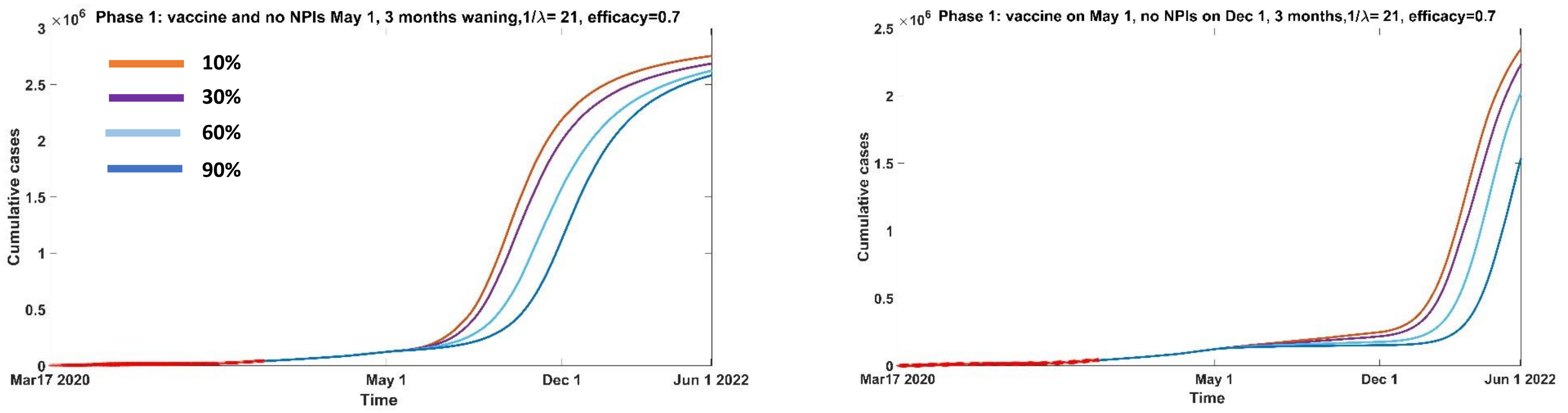


**A**

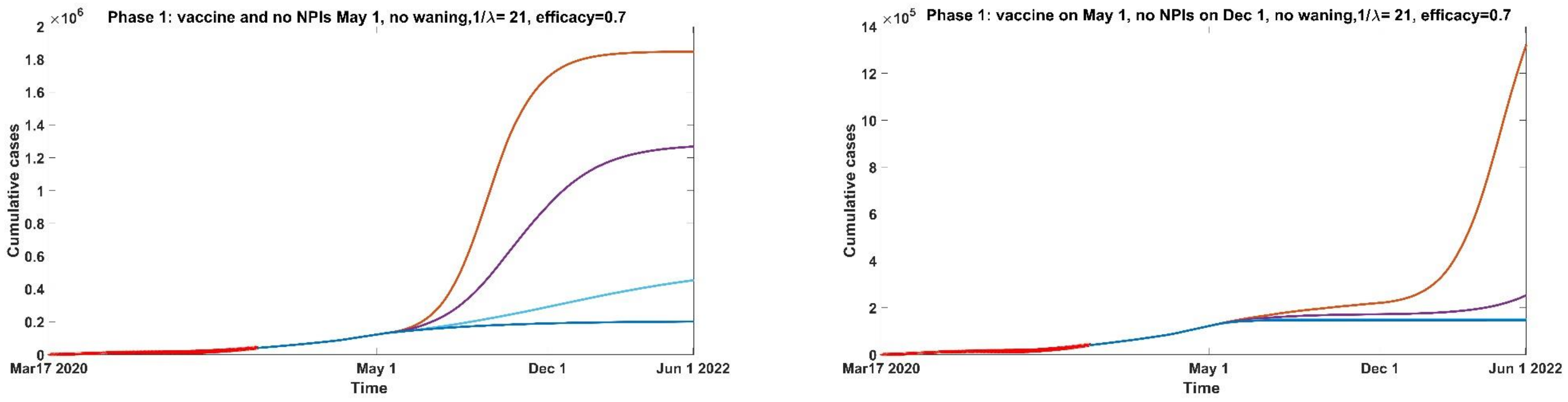


**B**

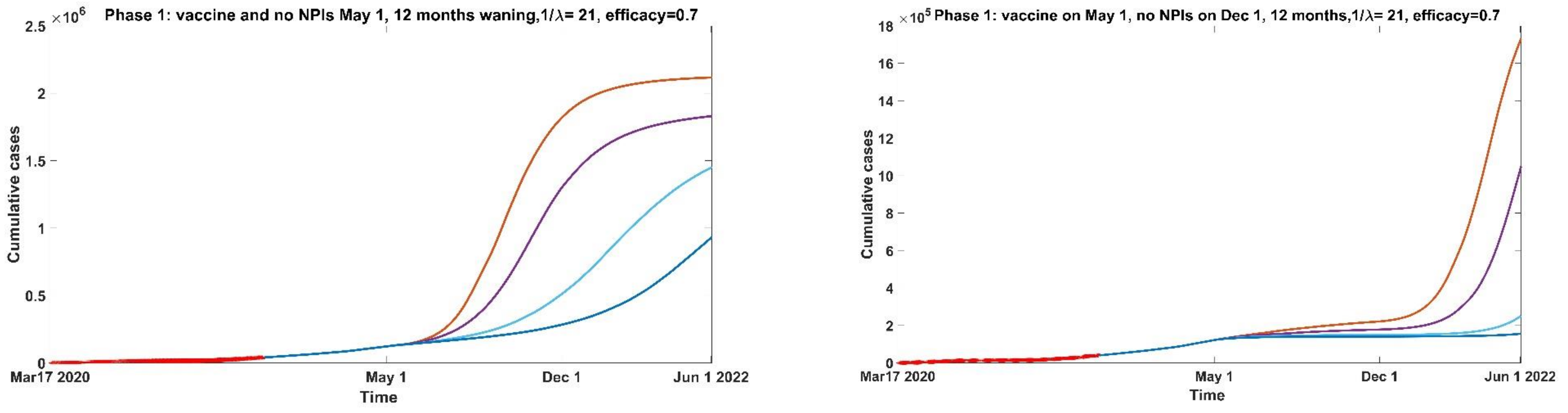


**C**

**Figure S3.1:** Cumulative cases from March 17, 2020 to June 2022 under Phase 1 when immunity (A) wanes over 3 months, (B) doesn't wane, (C) wanes over 12 months. Vaccine is introduced on May 1, 2021 (10%, orange, 30%, purple, 60% light blue, 90% blue) and distributed over 21 days. NPI's are lifted (highest number of contacts and probability of transmission) on December 1, 2021 (right panels) or in concomitance with vaccine (left panels), and vaccine is 70% efficient.

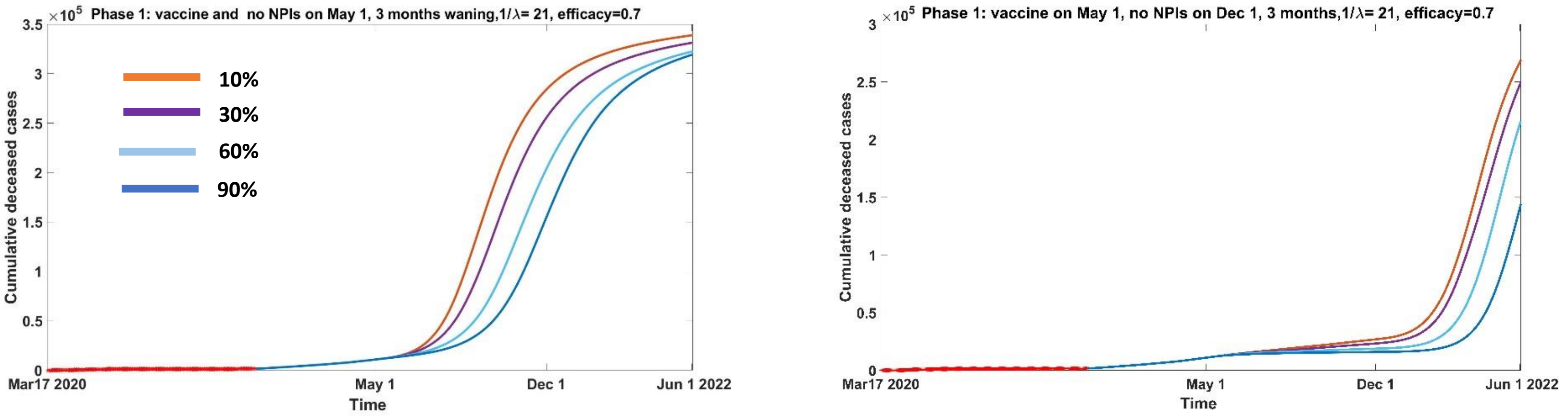


A

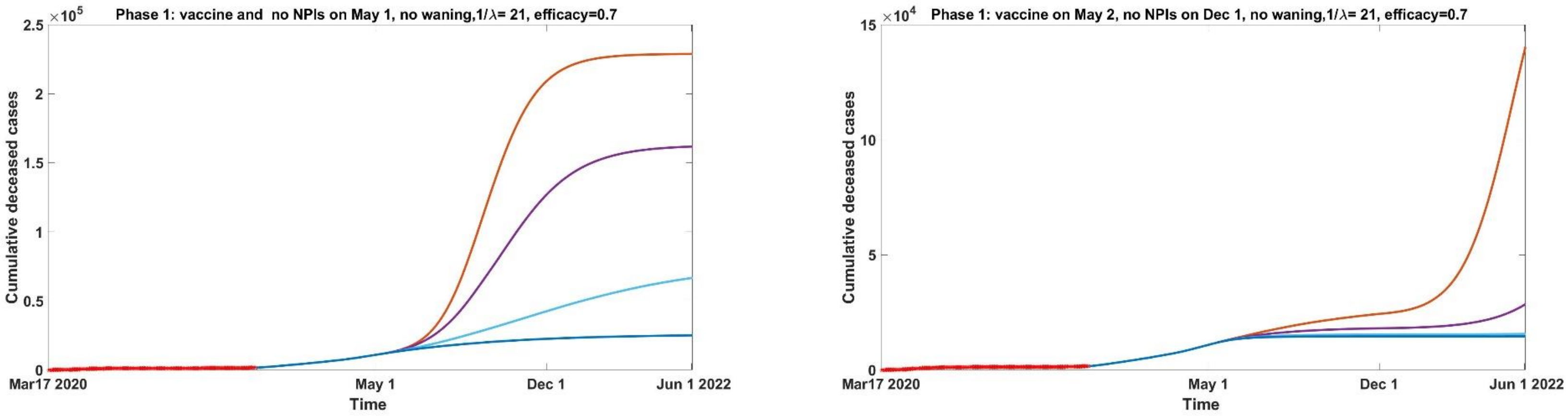


B

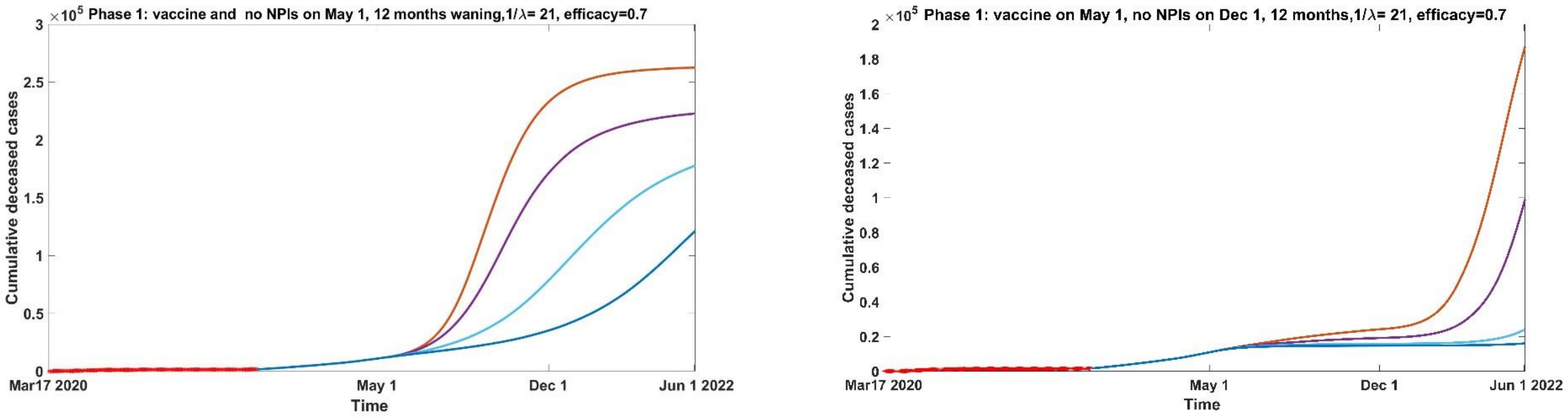


C

**Figure S3.2:** Cumulative deceased cases from March 17, 2020 to June 2022 under Phase 1 when immunity (A) wanes over 3 months, (B) doesn't wane, (C) wanes over 12 months. Vaccine is introduced on May 1, 2021 (10%, orange, 30%, purple, 60% light blue, 90% blue) and distributed over 21 days. NPI's are lifted (highest number of contacts and probability of transmission) on December 1, 2021 (right panels) or in concomitance with vaccine (left panels), and vaccine is 70% efficient.

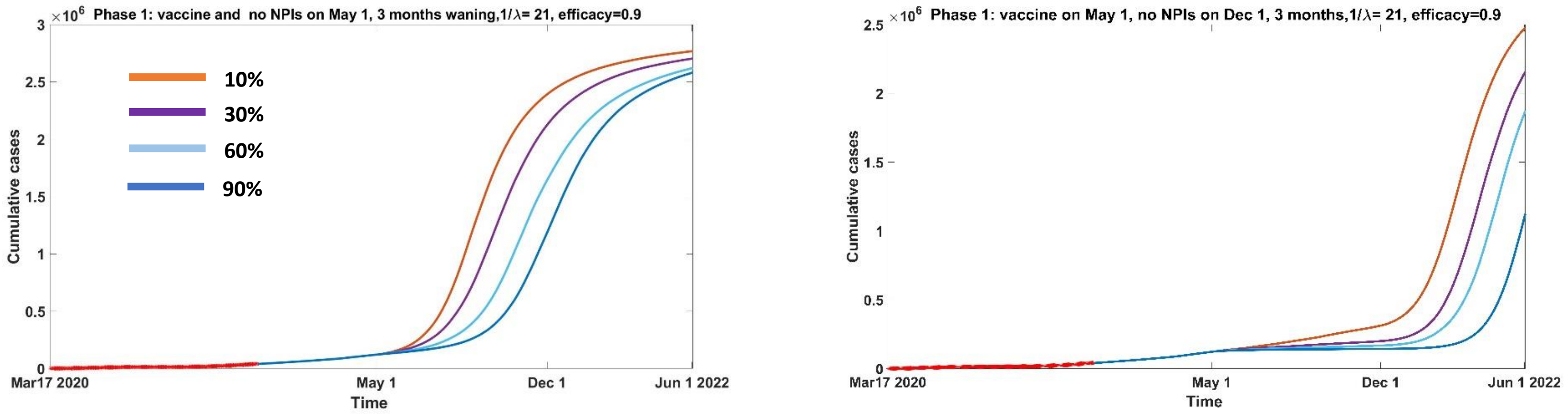


A

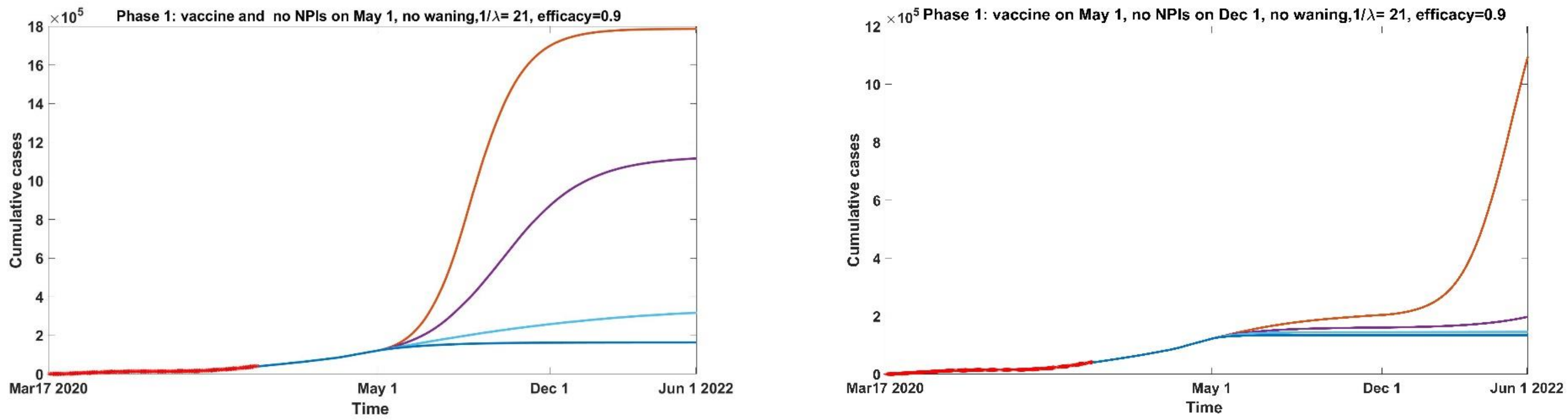


B

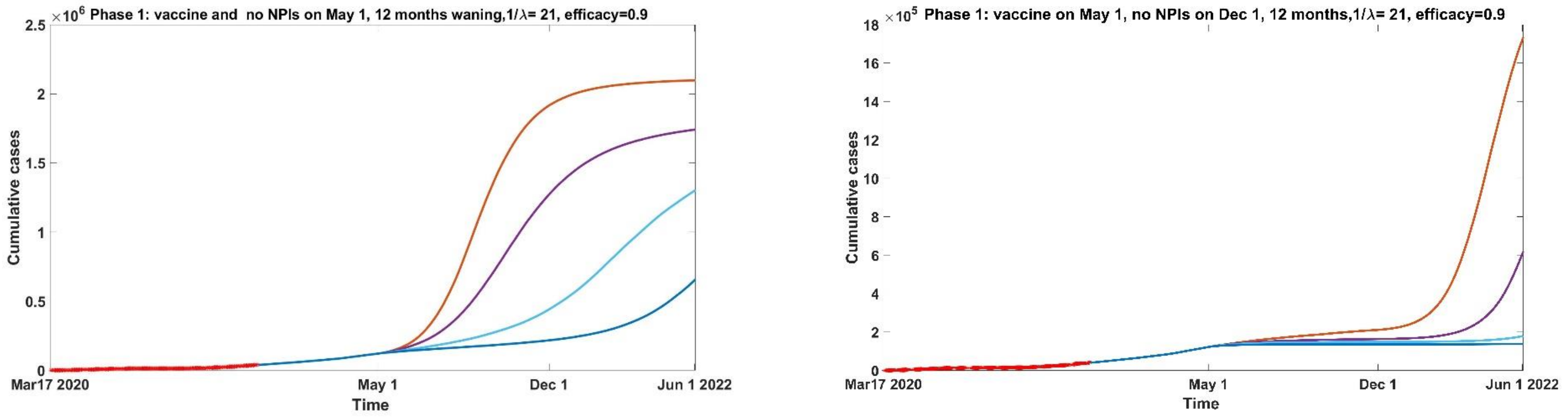


C

**Figure S3.3:** Cumulative cases from March 17, 2020 to June 2022 under Phase 1 when immunity (A) wanes over 3 months, (B) doesn't wane, (C) wanes over 12 months. Vaccine is introduced on May 1, 2021 (10%, orange, 30%, purple, 60% light blue, 90% blue) and distributed over 21 days. NPI's are lifted (highest number of contacts and probability of transmission) on December 1, 2021 (right panels) or in concomitance with vaccine (left panels), and vaccine is 90% efficient.

#### Phase 2

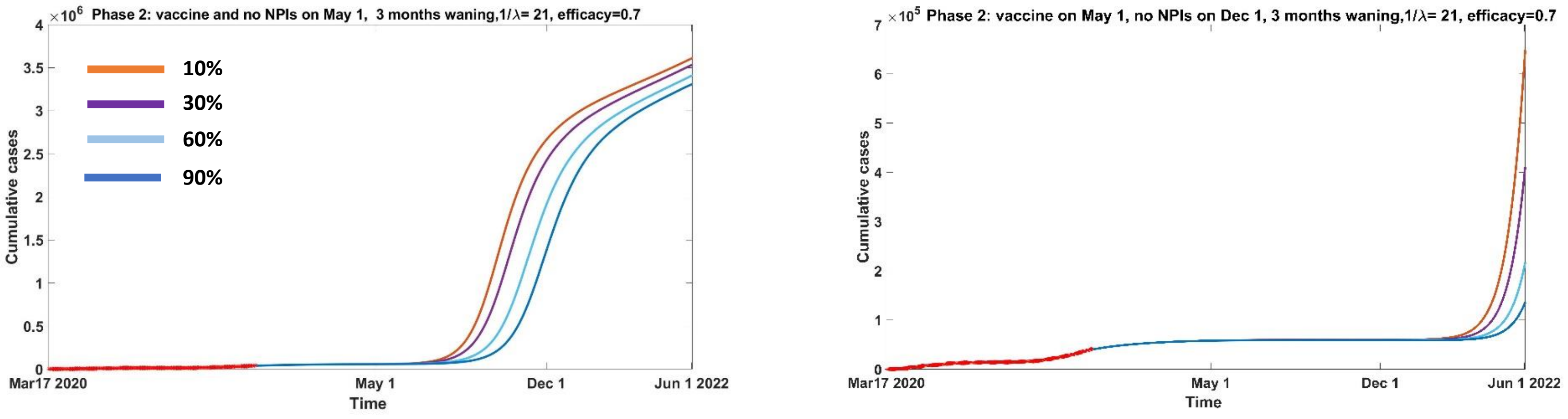


**A**

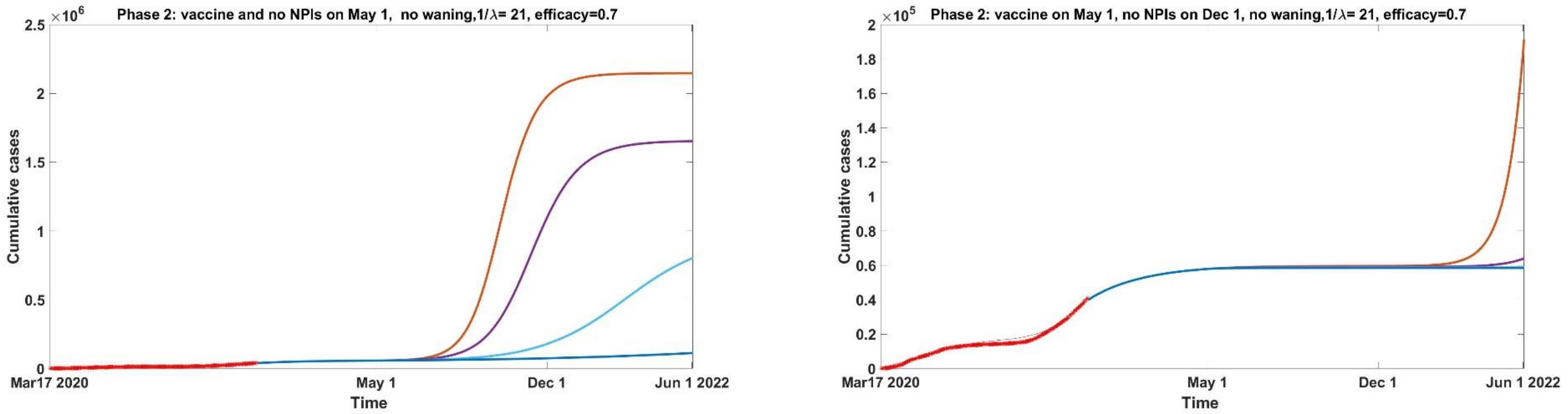


**B**

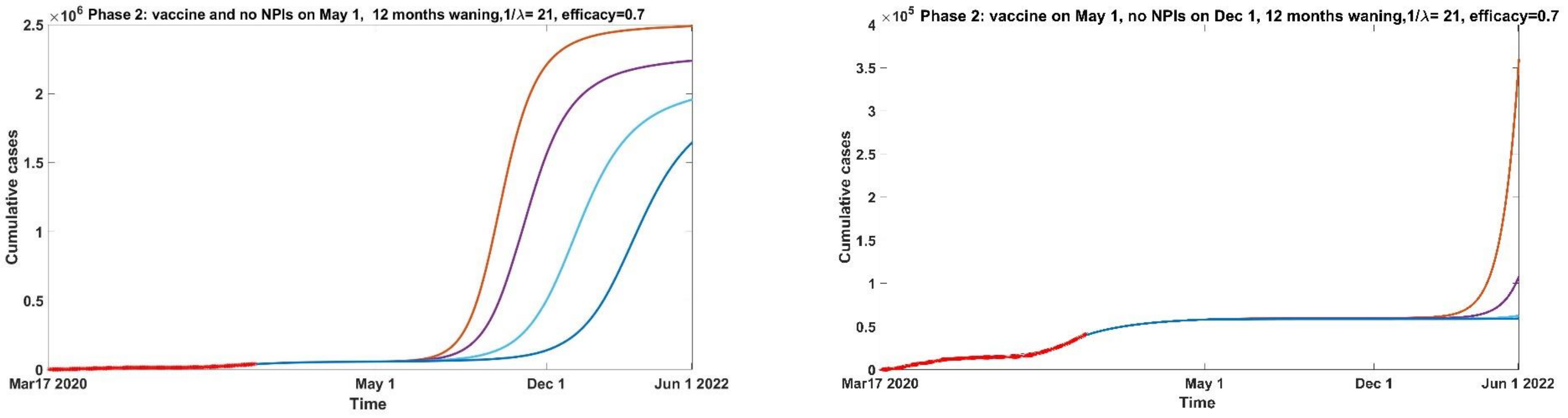


**C**

**Figure S3.4:** Cumulative cases from March 17, 2020 to June 2022 under Phase 2 when immunity (A) wanes over 3 months, (B) doesn't wane, (C) wanes over 12 months. Vaccine is introduced on May 1, 2021 (10%, orange, 30%, purple, 60% light blue, 90% blue) and distributed over 21 days. NPI's are lifted (highest number of contacts and probability of transmission) on December 1, 2021 (right panels) or in concomitance with vaccine (left panels), and vaccine is 70% efficient.

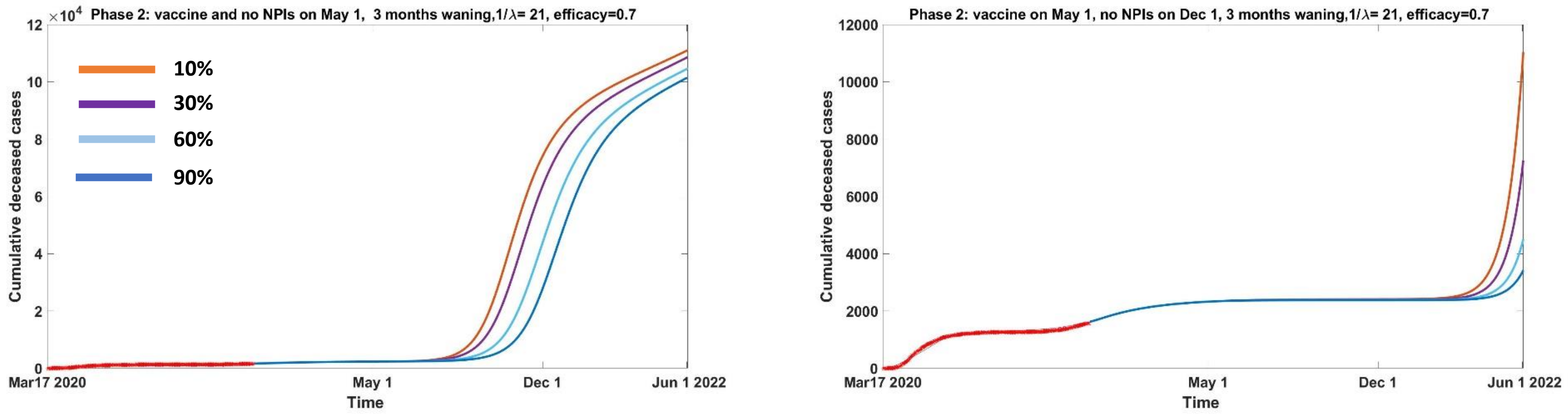


**A**

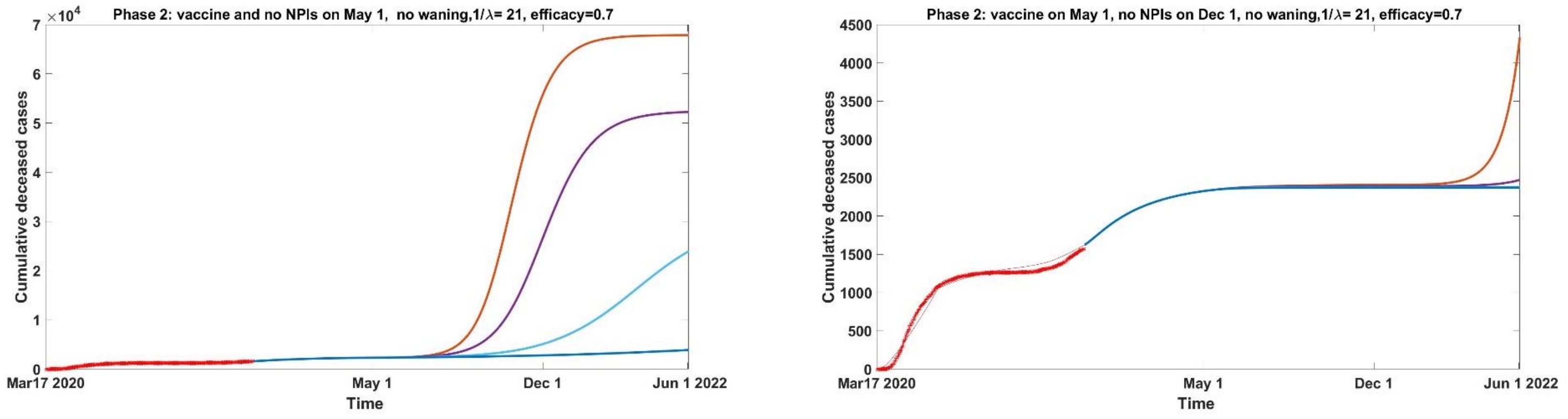


**B**

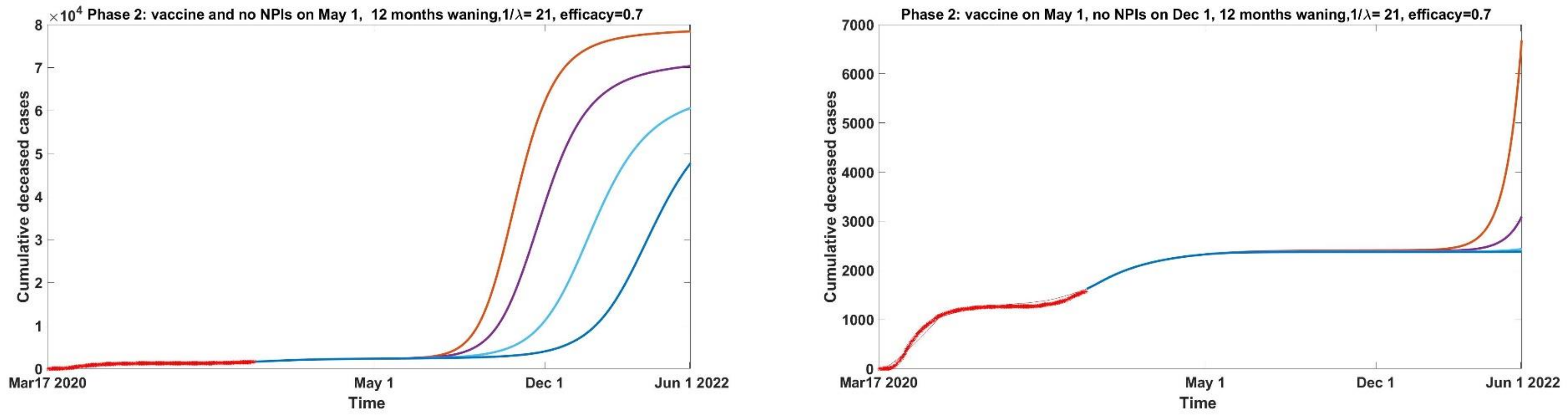


**C**

**Figure S3.5:** Cumulative deceased cases from March 17, 2020 to June 2022 under Phase 2 when immunity (A) wanes over 3 months, (B) doesn't wane, (C) wanes over 12 months. Vaccine is introduced on May 1, 2021 (10%, orange, 30%, purple, 60% light blue, 90% blue) and distributed over 21 days. NPI's are lifted (highest number of contacts and probability of transmission) on December 1, 2021 (right panels) or in concomitance with vaccine (left panels), and vaccine is 70% efficient.

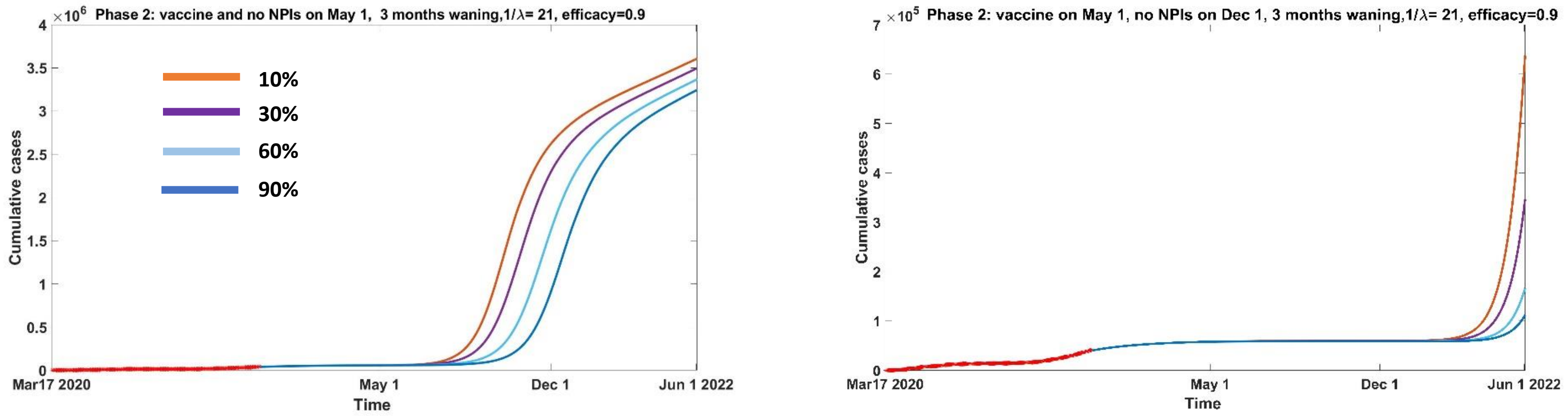


A

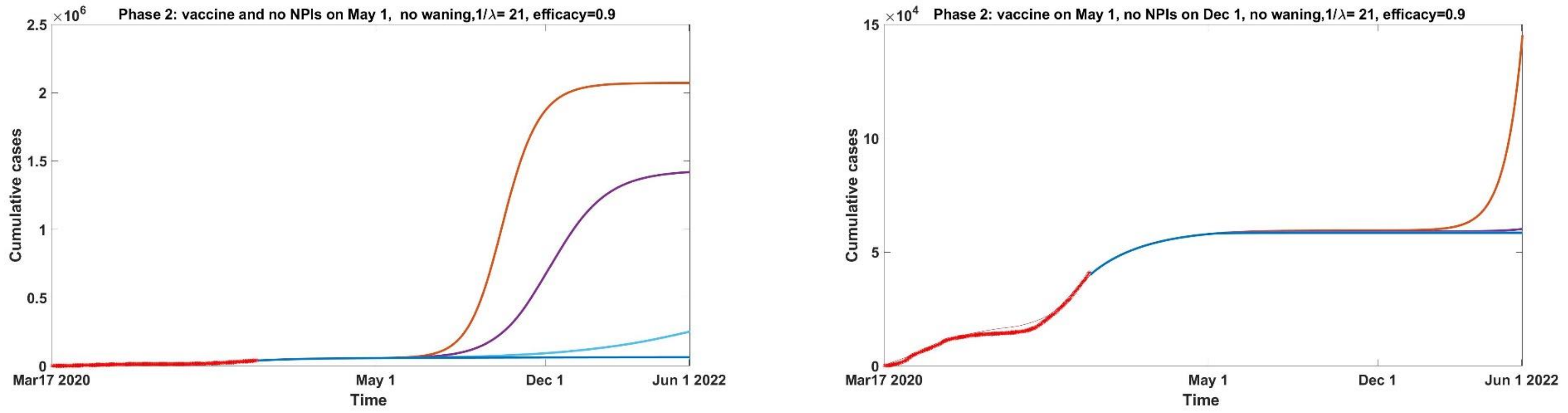


B

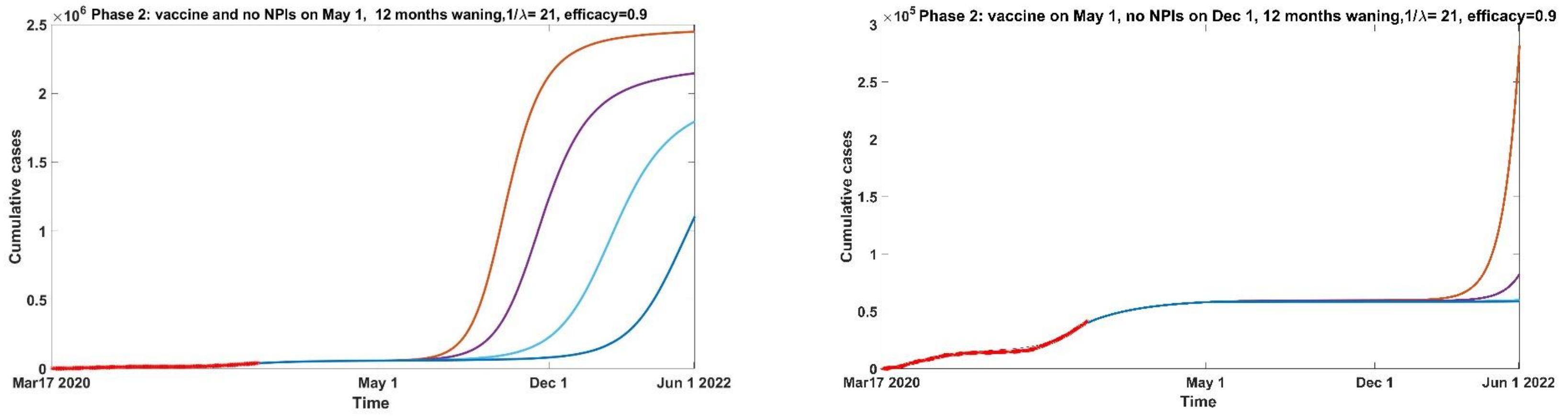


C

**Figure S3.6:** Cumulative cases from March 17, 2020 to June 2022 under Phase 2 when immunity (A) wanes over 3 months, (B) doesn't wane, (C) wanes over 12 months. Vaccine is introduced on May 1, 2021 (10%, orange, 30%, purple, 60% light blue, 90% blue) and distributed over 21 days. NPI's are lifted (highest number of contacts and probability of transmission) on December 1, 2021 (right panels) or in concomitance with vaccine (left panels), and vaccine is 90% efficient.

#### Phase 3

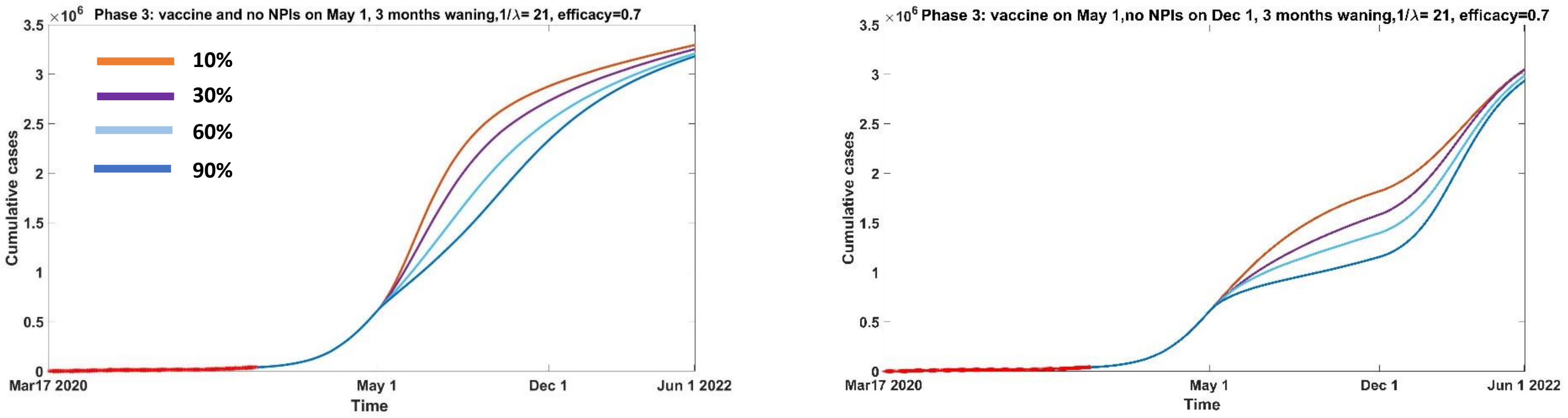


**A**

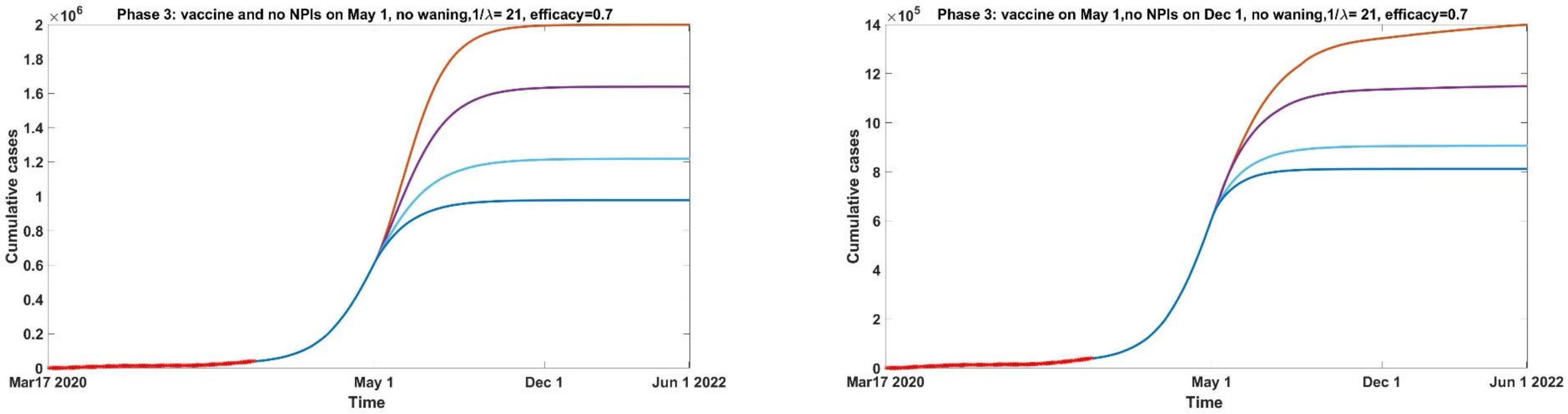


**B**

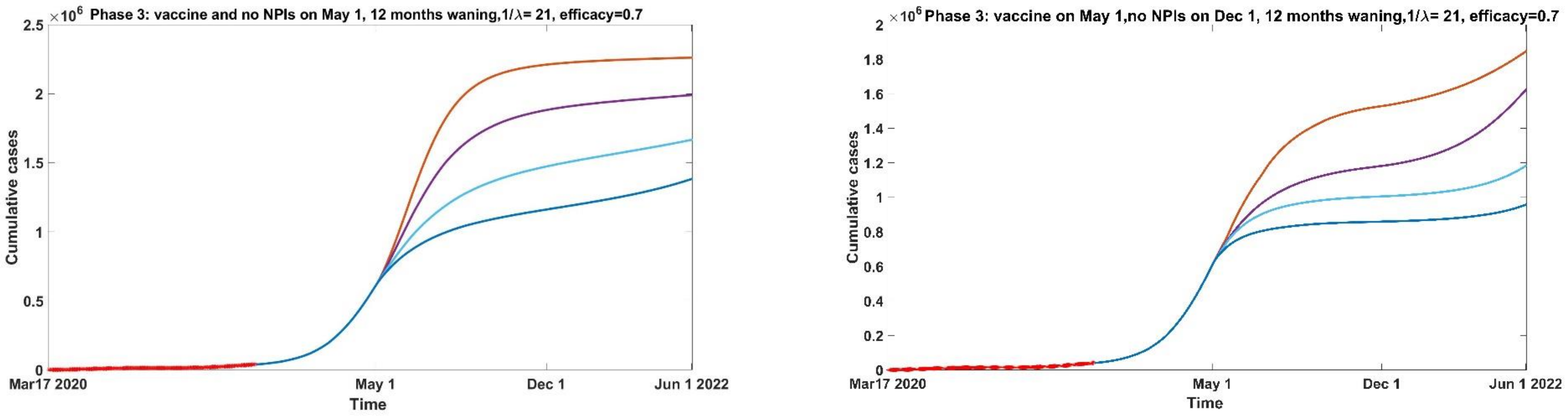


**C**

**Figure S3.7:** Cumulative cases from March 17, 2020 to June 2022 under Phase 3 when immunity (A) wanes over 3 months, (B) doesn't wane, (C) wanes over 12 months. Vaccine is introduced on May 1, 2021 (10%, orange, 30%, purple, 60% light blue, 90% blue) and distributed over 21 days. NPI's are lifted (highest number of contacts and probability of transmission) on December 1, 2021 (right panels) or in concomitance with vaccine (left panels), and vaccine is 70% efficient.

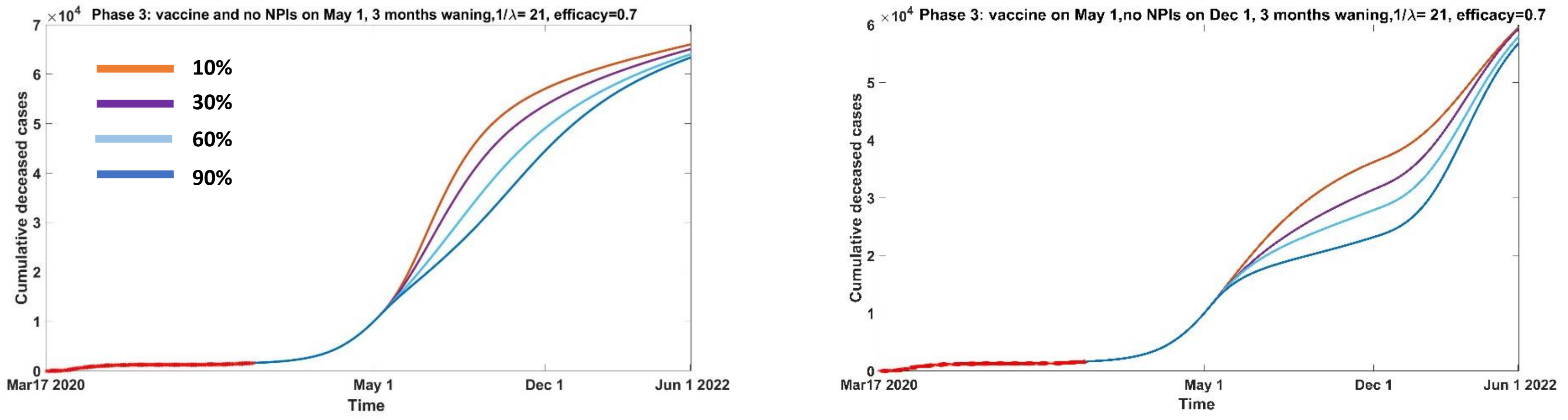


A

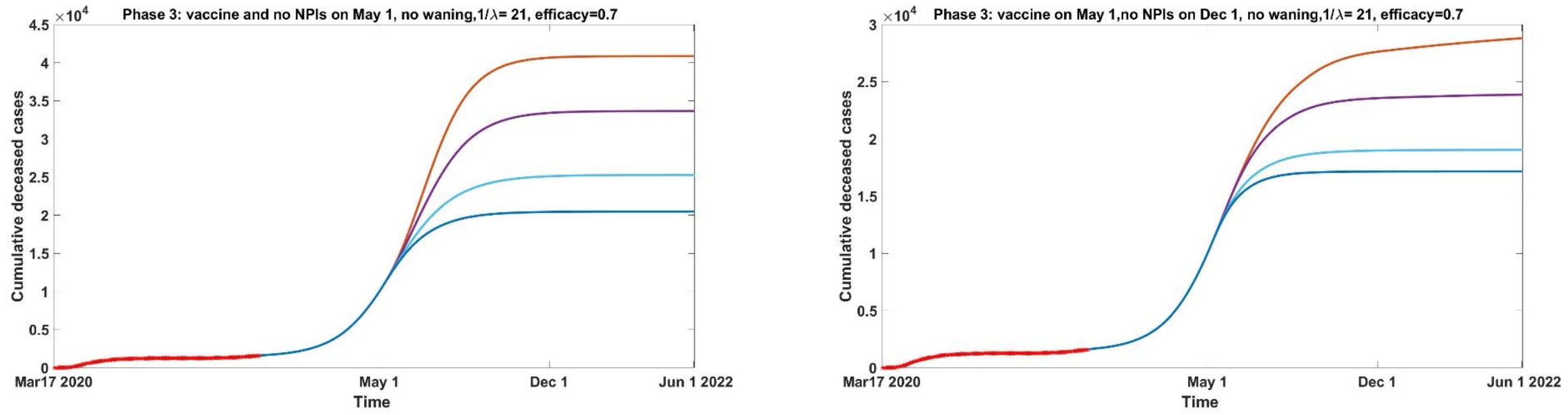


B

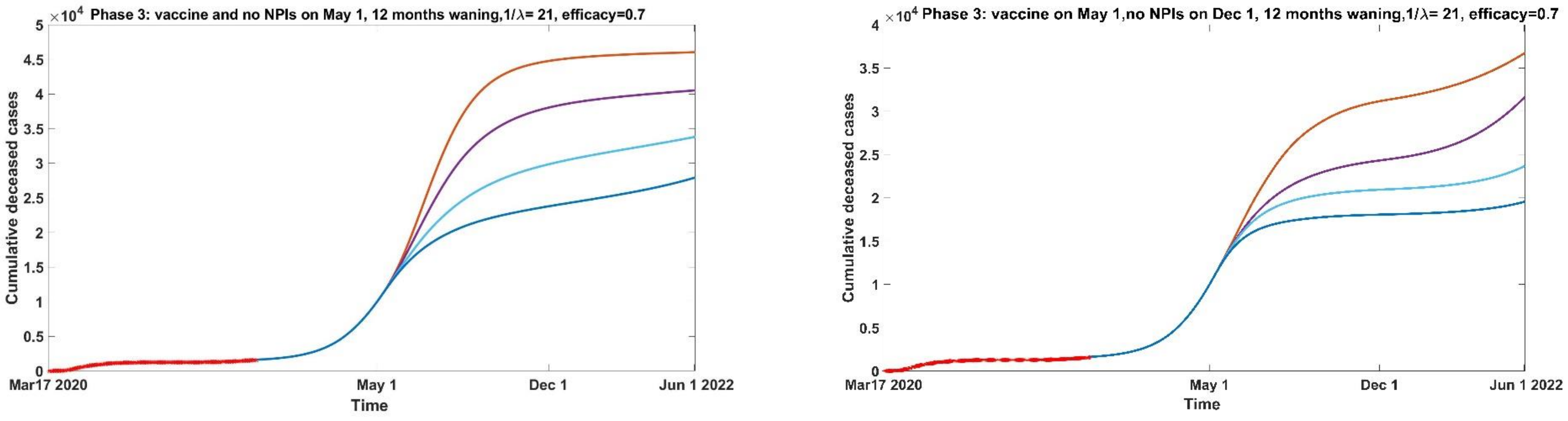


C

**Figure S3.8:** Cumulative deceased cases from March 17, 2020 to June 2022 under Phase 3 when immunity (A) wanes over 3 months, (B) doesn't wane, (C) wanes over 12 months. Vaccine is introduced on May 1, 2021 (10%, orange, 30%, purple, 60% light blue, 90% blue) and distributed over 21 days. NPI's are lifted (highest number of contacts and probability of transmission) on December 1, 2021 (right panels) or in concomitance with vaccine (left panels), and vaccine is 70% efficient.

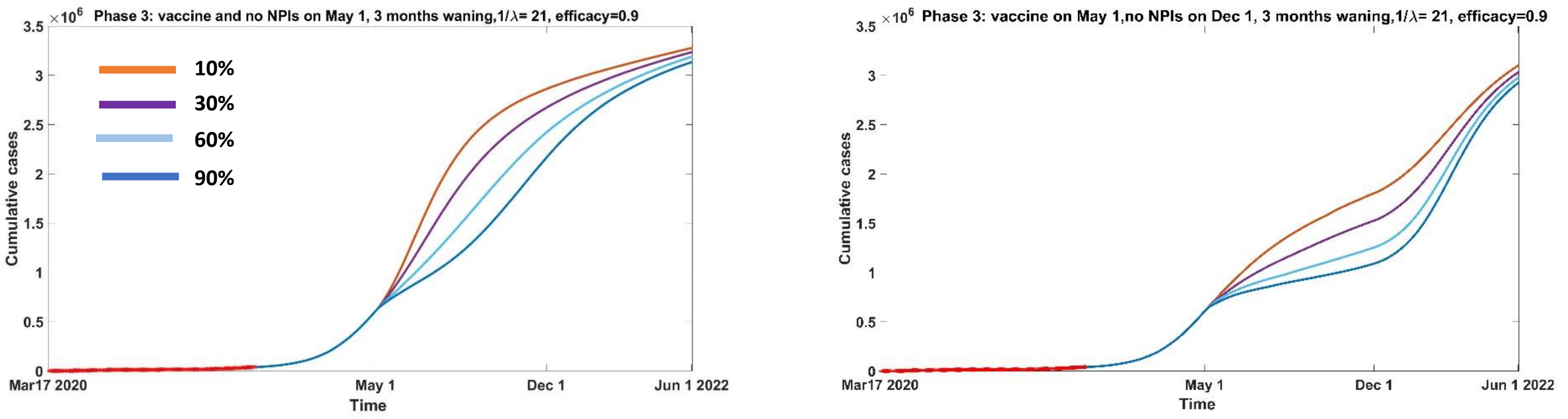


A

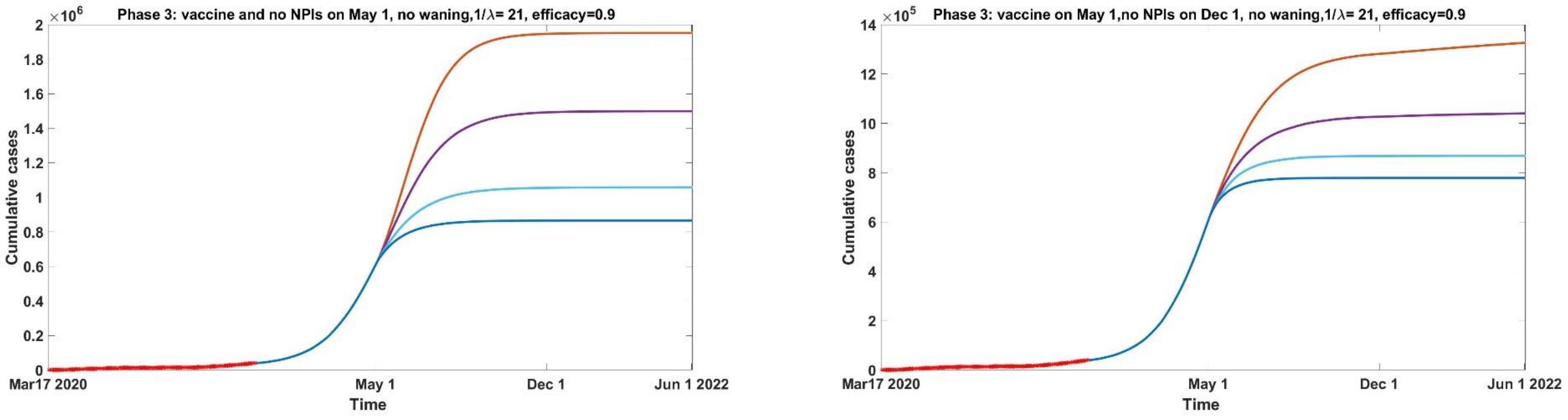


B

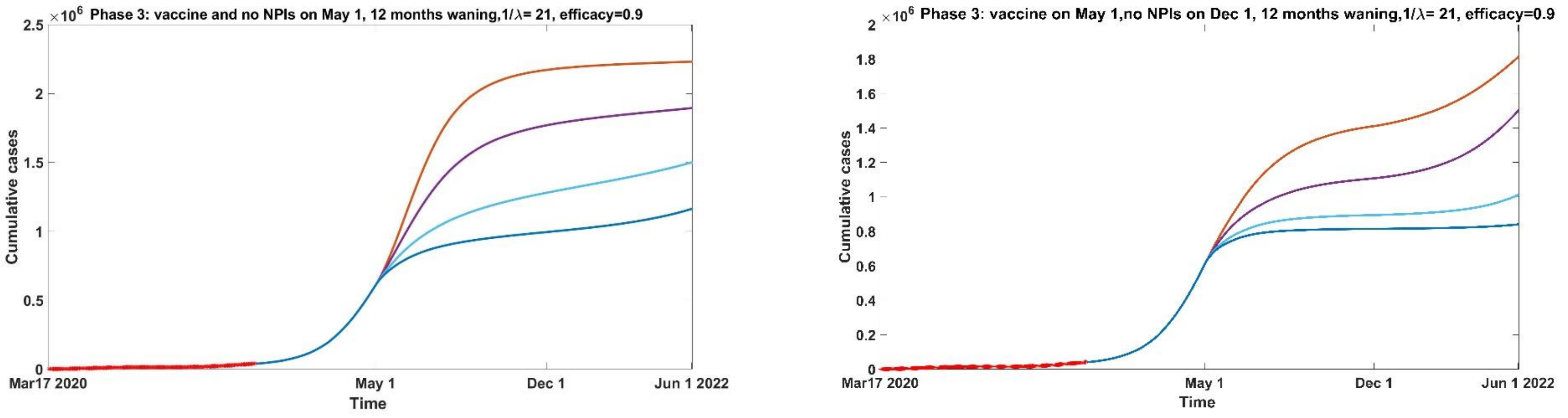


C

**Figure S3.9:** Cumulative cases from March 17, 2020 to June 2022 under Phase 3 when immunity (A) wanes over 3 months, (B) doesn't wane, (C) wanes over 12 months. Vaccine is introduced on May 1, 2021 (10%, orange, 30%, purple, 60% light blue, 90% blue) and distributed over 21 days. NPI's are lifted (highest number of contacts and probability of transmission) on December 1, 2021 (right panels) or in concomitance with vaccine (left panels), and vaccine is 90% efficient.

#### Phase 4

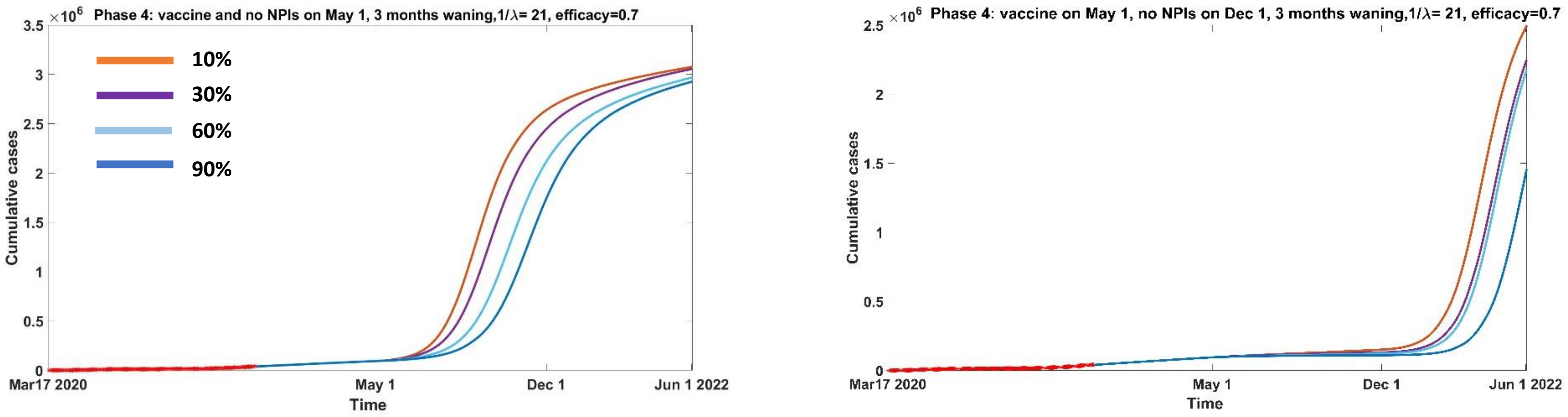


**A**

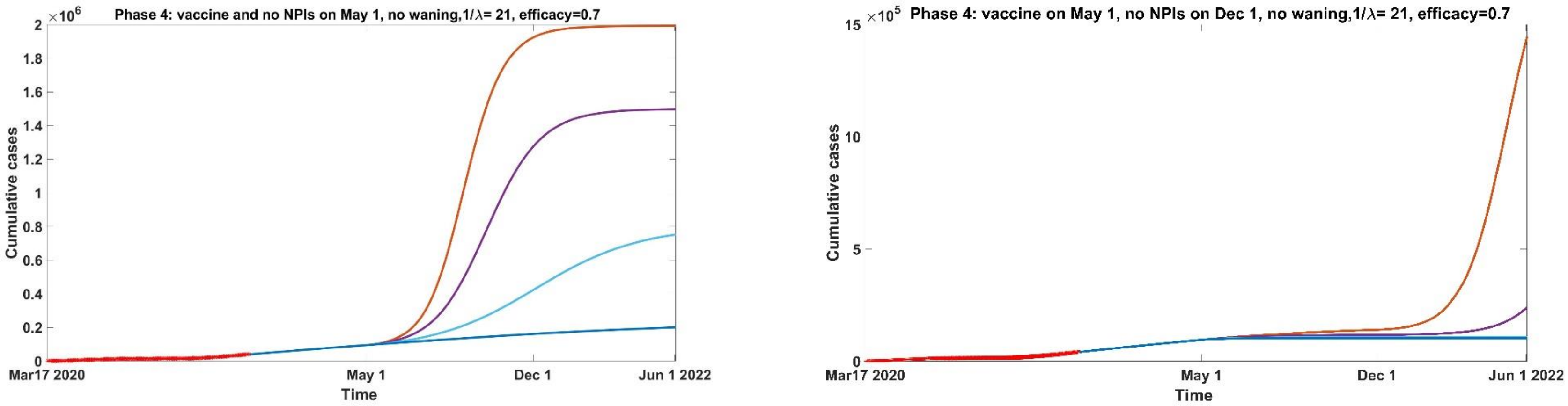


**B**

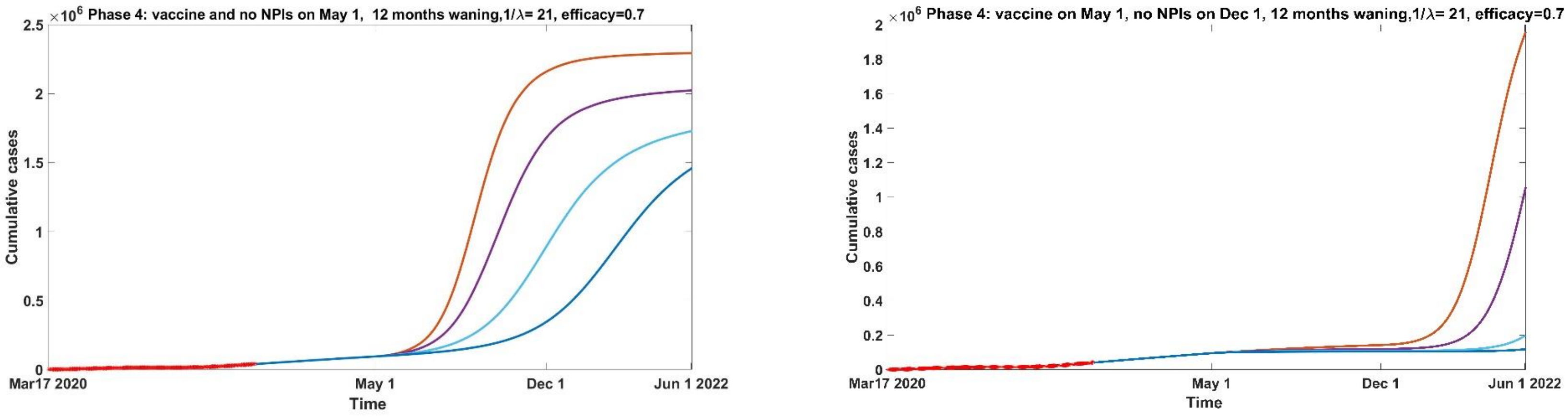


**C**

**Figure S3.10:** Cumulative cases from March 17, 2020 to June 2022 under Phase 4 when immunity (A) wanes over 3 months, (B) doesn't wane, (C) wanes over 12 months. Vaccine is introduced on May 1, 2021 (10%, orange, 30%, purple, 60% light blue, 90% blue) and distributed over 21 days. NPI's are lifted (highest number of contacts and probability of transmission) on December 1, 2021 (right panels) or in concomitance with vaccine (left panels), and vaccine is 70% efficient.

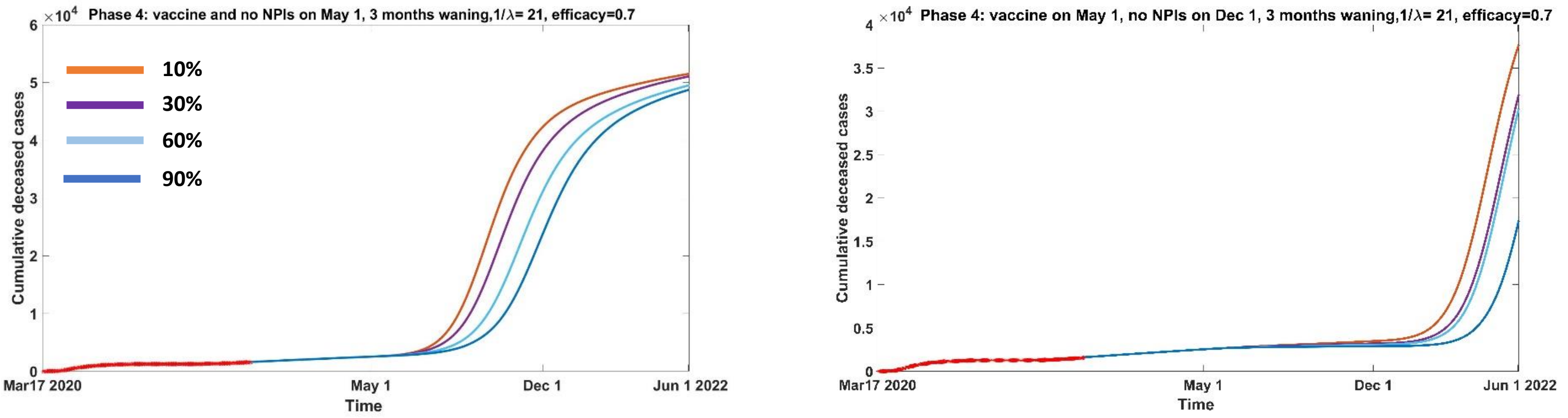


**A**

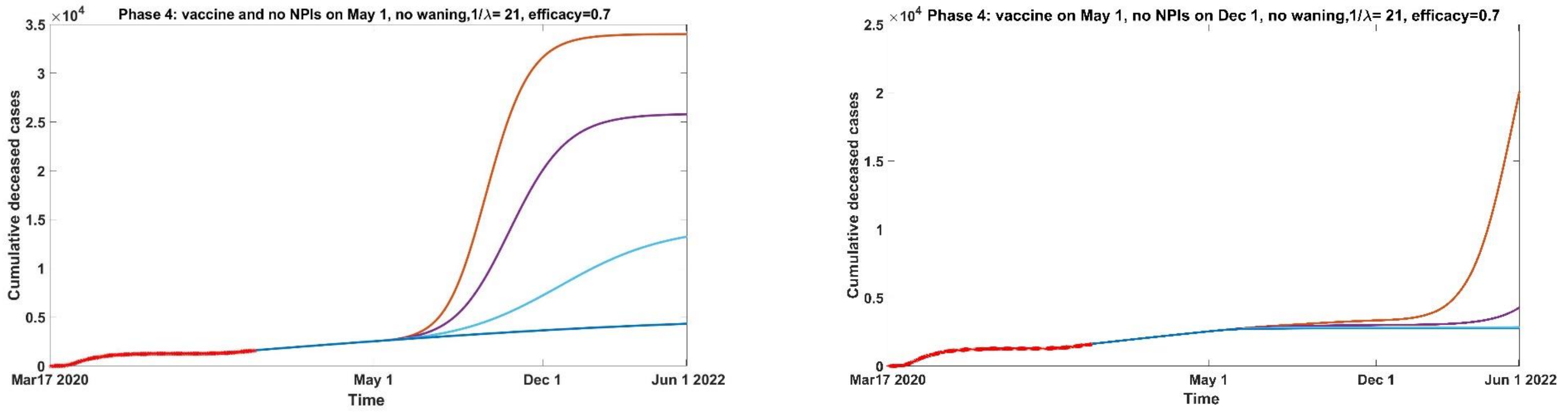


**B**

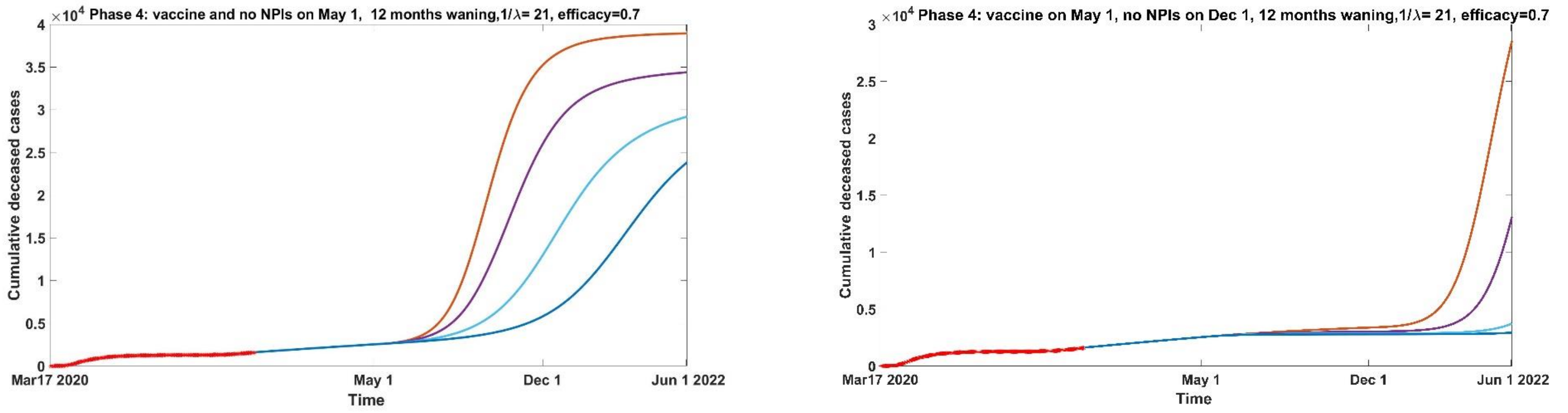


**C**

**Figure S3.11:** Cumulative deceased cases from March 17, 2020 to June 2022 under Phase 4 when immunity (A) wanes over 3 months, (B) doesn't wane, (C) wanes over 12 months. Vaccine is introduced on May 1, 2021 (10%, orange, 30%, purple, 60% light blue, 90% blue) and distributed over 21 days. NPI's are lifted (highest number of contacts and probability of transmission) on December 1, 2021 (right panels) or in concomitance with vaccine (left panels), and vaccine is 70% efficient.

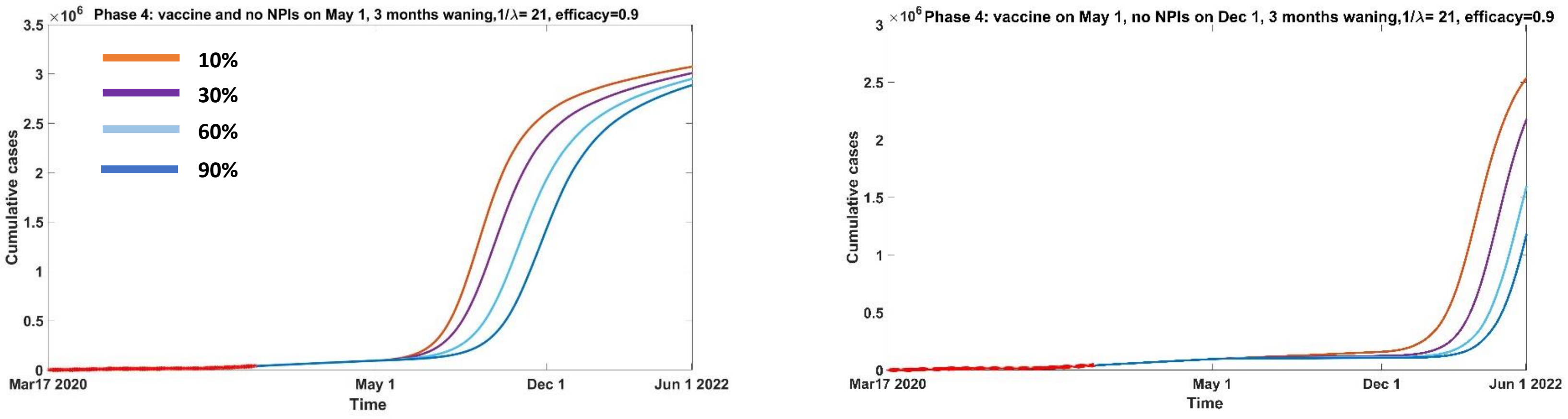


A

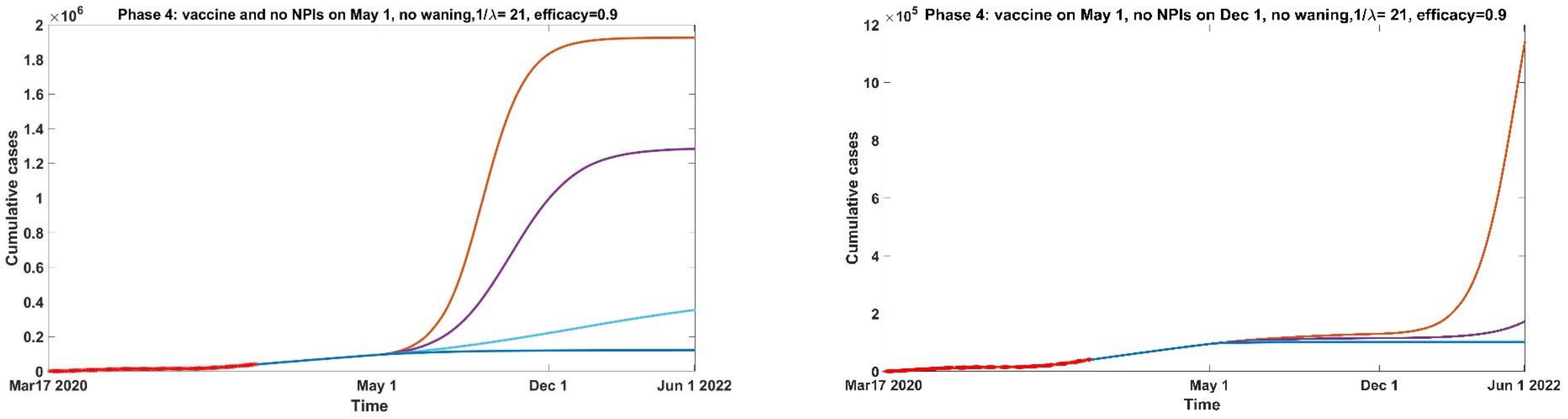


B

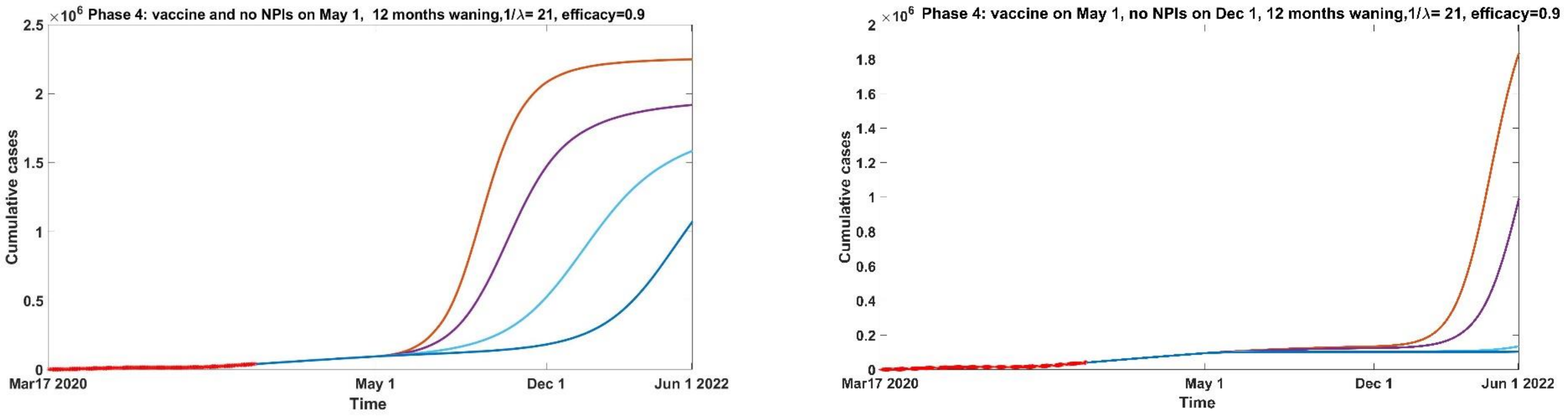


C

**Figure S3.12:** Cumulative cases from March 17, 2020 to June 2022 under Phase 4 when immunity (A) wanes over 3 months, (B) doesn't wane, (C) wanes over 12 months. Vaccine is introduced on May 1, 2021 (10%, orange, 30%, purple, 60% light blue, 90% blue) and distributed over 21 days. NPI's are lifted (highest number of contacts and probability of transmission) on December 1, 2021 (right panels) or in concomitance with vaccine (left panels), and vaccine is 90% efficient.

**References (Supplementary Materials)**

[7]City of Toronto https://www.toronto.ca/home/covid-19/covid-19-latest-city-of-toronto-news/covid-19-status-of-cases-in-toronto/

[8] Government of Ontario https://covid-19.ontario.ca/data

[9] City of Toronto https://www.toronto.ca/city-government/data-research-maps/toronto-at-a-glance/

[10] Diekmann, O.; Heesterbeek, J. A. P.; Metz, J. A. J. (1990). "On the definition and the computation of the basic reproduction ratio $R_0$ in models for infectious diseases in heterogeneous populations". Journal of Mathematical Biology . **28** (4): 365–382. doi:10.1007/BF00178324. hdl: 1874/8051 . PMID 2117040.

[11] Heffernan J.M, Smith R.J, Wahl L.M, 2005, Perspectives on the basic reproductive ratio J. R. Soc. Interface.2281–293 http://doi.org/10.1098/rsif.2005.0042

[12] Van den Driessche, P.; Watmough, J. (2002). "Reproduction numbers and sub-threshold endemic equilibria for compartmental models of disease transmission". Mathematical Biosciences. **180** (1–2): 29–48. doi:10.1016/S0025-5564(02)00108-6. PMID 12387915
